# The novel pathway phenotype “major neurocognitive psychosis” is validated as a distinct class through the analysis of immune-linked neurotoxicity biomarkers and neurocognitive deficits

**DOI:** 10.1101/2024.04.17.24305941

**Authors:** Petar Popov, Chen Chen, Hussein Kadhem Al-Hakeim, Ali Fattah Al-Musawi, Arafat Hussein Al-Dujaili, Drozdstoy Stoyanov, Michael Maes

**Affiliations:** Department of Psychiatry, Medical University of Plovdiv, Plovdiv, Bulgaria; Sichuan Provincial Center for Mental Health, Sichuan Provincial People’s Hospital, School of Medicine, University of Electronic Science and Technology of China, Chengdu 610072, China; Key Laboratory of Psychosomatic Medicine, Chinese Academy of Medical Sciences, Chengdu, 610072, China; Department of Chemistry, College of Science, University of Kufa, Iraq; Department of Clinical Pharmacy and Laboratory Sciences, College of Pharmacy, University of Al-Kafeel, Najaf, Iraq; Department of Psychiatry, Faculty of Medicine, University of Kufa, Najaf, Iraq; Research Institute, Medical University Plovdiv, Plovdiv, Bulgaria; Research and Innovation Program for the Development of MU - PLOVDIV–(SRIPD-MUP)”, Creation of a network of research higher schools, National plan for recovery and sustainability, European Union – NextGenerationEU; Department of Psychiatry, Faculty of Medicine, Chulalongkorn University, Bangkok, Thailand; Kyung Hee University, 26 Kyungheedae-ro, Dongdaemun-gu, Seoul 02447, Korea

**Keywords:** schizophrenia, neuroimmune, biomarkers, inflammation, immune-associated neurotoxicity

## Abstract

**Background:** Using machine learning methods based on neurocognitive deficits and neuroimmune biomarkers, two distinct classes were discovered within schizophrenia patient samples. The first, major neurocognitive psychosis (MNP) was characterized by cognitive deficits in executive functions and memory, higher prevalence of psychomotor retardation, formal thought disorders, mannerisms, psychosis, hostility, excitation, and negative symptoms, and diverse neuroimmune aberrations. Simple neurocognitive psychosis (SNP) was the less severe phenotype.

**Aims:** The study comprised a sample of 40 healthy controls and 90 individuals diagnosed with schizophrenia, divided into MNP and SNP based on previously determined criteria. Soft Independent Modelling of Class Analogy (SIMCA) was performed using neurocognitive test results and measurements of serum M1 macrophage cytokines, IL-17, IL-21, IL-22, and IL-23 as discriminatory/modelling variables. The model-to-model distances between controls and MNP+SNP and between MNP and SNP were computed, and the top discriminatory variables were established.

**Results:** A notable SIMCA distance of 146.1682 was observed between MNP+SNP and the control group; the top-3 discriminatory variables were lowered motor speed, an activated T helper-17 axis, and lowered working memory. This study successfully differentiated MNP from SNP yielding a SIMCA distance of 19.3. M1 macrophage activation, lowered verbal fluency, and executive functions were the prominent features of MNP versus SNP.

**Discussion:** Based on neurocognitive assessments and the immune-linked neurotoxic IL-6/IL-23/Th-17 axis, we found that MNP and SNP are qualitatively distinct classes. Future biomarker research should always examine biomarkers in the MNP versus SNP phenotypes, rather than in the combined MNP + SNP or schizophrenia group.

## Introduction

Approximately 24 million individuals, or 1 in 300 persons (0.32%) globally, are afflicted with schizophrenia [1]. A combination of physical, genetic, psychological, and environmental factors may increase an individual’s susceptibility to schizophrenia, according to research [2, 3]. Schizophrenia is one of the most debilitating illnesses and individuals diagnosed with this illness experience profound distress across various critical domains, including social, familial, and personal spheres, among others [4]. Furthermore, in comparison to the general population, individuals afflicted with this disorder experience a reduced life expectancy as a result of complications including metabolic, cardiovascular, and infectious ailments [5].

Dementia praecox, a term originally proposed by Kraepelin in 1887, is defined as a progressive deterioration of cognitive functions accompanied by a “defect” and “productive” symptoms concurrently [6–11]. Subsequently, in 1911, Bleuler introduced the term “schizophrenia” to denote the cognitive disarray and psychiatric condition that individuals afflicted with schizophrenia experience. Bleuler (2011) offered a critique of the terminology “dementia praecox,” highlighting the fact that cognitive decline is not an inevitable outcome of the ailment [12, 13]. He regarded schizophrenia as consisting of two symptom dimensions: a core containing primary symptoms (abnormal thought processes, aberrant emotion, autistic behaviors, and ambivalence), and accessory symptoms (social withdrawal, delusions, hallucinations, and diminished desire) [14–16]. According to Schneider (1959), the primary symptoms of schizophrenia include delusions, hallucinations (particularly somatic and auditory), thought withdrawal and interruptions, and thought dissemination [14, 17, 18]. Snezhnevsky defined the “defect” as a decline in social engagement and mental capacities, diminished energy potential and mental fatigue, social withdrawal, mental marasmus, dulled affect, personality regression, and neurocognitive decline [19, 20]. Unfortunately, the American categorization system, considered the gold standard in schizophrenia research, disregards this most valuable European and Russian information, and focuses only on one diagnostic category of schizophrenia (DSM-IV to DSM-5).

Maes and his research groups [21, 22] using machine learning methods a) defined the “defect” as a latent construct score extracted from various symptom dimensions, cognitive test results comprising memory and executive functions, disabilities, and health-related quality of life data (physical, psychological, and social domains); and b) discovered two distinct classes of schizophrenia patients [21, 23]. One significant distinction between the two subgroups is that the most severe subgroup exhibits a higher prevalence of psychomotor retardation, formal thought disorders, mannerisms, psychosis, hostility, excitation, and negative symptoms such as blunted affect, alogia, asociality, anhedonia, and avolition [21]. Furthermore, this particular subgroup is distinguished by heightened disabilities, diminished health-related quality of life, and particularly anomalies in neurocognitive processes, such as executive functions and memory [24, 25].

Kanchanatawan et al. [23] suggested substituting the stigmatizing term “schizophrenia” with “neurocognitive psychosis” and “major neurocognitive psychosis (MNP)” to designate the most severe subtype, while “simple neurocognitive psychosis (SNP)” be used to designate the less severe subtype. Nonetheless, specific supervised machine learning (Soft Independent Modelling of Class Analogy or SIMCA) determined that MNP and SNP are qualitatively distinct groups on the basis of biomarker studies and neurocognitive test results [21]. First, it is important to note that MNP is characterized by significant cognitive deficits including attention, learning, memory, decision-making, perception, recognition, and sensory input [26]. Significantly, a latent vector denoted as generalized cognitive decline (G-CoDe) could be extracted from the various neurocognitive test results [26]. The G-CoDe score was significantly lowered in MNP as compared with SNP and controls indicating that the “defect” or “cognitive deficit” is most pronounced in MNP [26]. Second, MNP is distinguished by a multitude of neuroimmune pathways in comparison to healthy controls and SNP [21]. In general, the observed pathways suggest heightened immune-associated neurotoxicity as indicated by elevated concentrations of neurotoxic cytokines and chemokines, specifically interleukin-1β, CCL11, and tumor necrosis factor (TNF)-α, as well as neurotoxic tryptophan catabolites (TRYCATs) [21]. Additionally, there is an increased load of lipopolysaccharides (LPS) in the peripheral bloodstream and biomarkers of oxidative and nitrosative stress [21]. Additional notable characteristics of MNP include the suppression of protective outcome pathways, such as antioxidant enzyme activities and antioxidant defenses, as well as natural IgM-mediated autoimmune responses [21].

Furthermore, MNP is differentiated from SNP by an activated neurotoxic signaling pathway, namely the IL-6/IL-23/T-helper (Th)-17 axis. Indeed, Al-Hakeim et al. (2022) identified a latent vector score comprised of IL-6, IL-17, IL-21, IL-22, and IL-23 that exhibited a significantly higher value in MNP compared to SNP [24, 25]. This score was predictive of cognitive deficits and the severity of the phenotype of schizophrenia.

However, there is a lack of research investigating the possibility that MNP and SNP constitute distinct categories in terms of the IL-6/IL-23/Th-17 axis. Therefore, the purpose of this study was to investigate whether MNP and SNP constitute qualitatively distinct classes, as well as whether these two groups differ qualitatively from healthy controls. Nevertheless, traditional statistical tests lack the capability to distinguish whether diagnostic classes differ qualitatively. Furthermore, while prevalent supervised machine learning methods like neural networks, support vector machines, and linear discriminant analysis enable the classification of cases and controls, they do not possess the capability to assess qualitative distinctions among classes [27, 28]. Hence, employing SIMCA [27, 28], the present investigation was conducted to ascertain whether MNP constitutes a qualitatively distinct class as indicated by measurements of IL-6, IL-17, IL-21, IL-22, and IL-23 and neurocognitive tests [21].

## Subjects and Methods

### Participants

Schizophrenia patients were enrolled in the psychiatry unit of Al-Hakeem General Hospital in Najaf Governorate, Iraq, during the period of February to June 2021. The control group consisted of individuals who were either family members or acquaintances of personnel or patients and were recruited from the same catchment area (Najaf Governorate) as the patients. The study comprised a sample of 90 individuals diagnosed with schizophrenia and 40 healthy controls. All patients diagnosed with schizophrenia were assessed using the Mini-International Neuropsychiatric Interview (M.I.N.I.) in accordance with the DSM-IV-TR criteria. Following a minimum of 12 weeks of stability, the patients were categorized into two groups: those who presented with deficit schizophrenia and those who did not, as determined by the Schedule for the Deficit Syndrome (SDS) and as explained previously [21, 29]. In the twelve months prior to the diagnosis, MNP was identified as characterized by the presence of at least two of the following six symptoms with clinically significant severity: affect restriction, diminished emotional range, speech poverty, interest curtailment, diminished sense of purpose, and diminished social drive. Patients who presented with other axis-1 disorders as defined by the DSM-IV-TR were ineligible. These disorders comprised psycho-organic diseases, substance use disorders, schizoaffective disorder, severe major depression, bipolar disorder, and obsessive-compulsive disorder. All the controls lacked any lifetime diagnosis of disorders classified under DSM-IV-TR axis 1. The inclusion criteria for patients and controls were as follows: neurodegenerative or neuroinflammatory conditions (e.g., Alzheimer’s disease, Parkinson’s disease, multiple sclerosis, stroke); prior use of immunosuppressive or glucocorticoid therapy; (auto)immune disorders (e.g., rheumatoid arthritis, psoriasis, chronic obstructive pulmonary disease, inflammatory bowel disease, diabetes mellitus type 1. We excluded patients with tardive dyskinesia, akathisia, dystonia, neuroleptic malignant syndrome, and debilitating Parkinsonism (e.g., shuffling gait, bradykinesia). Women who were expectant or lactating were also excluded. Omitted were also individuals who took therapeutical doses of antioxidant or omega-3 supplements.

The research was conducted in accordance with Iraqi and international ethical and privacy standards. All participants and first-degree relatives of those with schizophrenia provided written informed consent prior to their involvement in this research. Legal representatives of these individuals include the mother, father, brother, spouse, or son. The research was approved by the ethics committee (IRB) of the College of Science, University of Kufa, Iraq (82/2020), in accordance with the International Guideline for the Protection of Human Research of the Declaration of Helsinki.

### Clinical assessments

To gather patient and control data, semi-structured interviews were administered by a senior psychiatrist who specialized in schizophrenia. The senior psychiatrist administered the Positive and Negative Syndrome Scale (PANSS) [30], the Scale for the Assessments of Negative Symptoms (SANS) [31], the Brief Psychiatric Rating Scale (BPRS) [32], and the Hamilton Depression (HAM-D) and Anxiety (HAM-A) Rating Scales [33, 34] on the same day. As previously described, we utilized BPRS, HAM-D, HAM-A, and PANSS items to construct z-unit weighted composite scores that indicated psychosis, hostility, excitement, mannerism, formal thought disorders (FTD), and psychomotor retardation (PMR). Negative symptoms were quantified using the total SANS, negative PANSS symptom sore.

A research psychologist conducted neuropsychological investigations utilizing the Brief Assessment of Cognition in Schizophrenia (BACS) [35] on the same day, maintaining a blind position about the clinical diagnosis. The List Learning Test (which measures verbal episodic memory), Digit Sequencing Task (which measures working memory), Category Instances (which measures semantic fluency), the Controlled Word Association (which measures letter fluency), Symbol Coding (which measures attention), the Tower of London (which measures executive functions), and the token motor task comprise the latter battery. The sum of all scores was calculated. Utilization of tobacco (TUD) was diagnosed in meeting the DSM-IV-TR criteria. The body mass index (BMI) was calculated utilizing the subsequent formula: body weight (kg) divided by length (m^2^).

### Assays

Early in the morning, fasting venous blood samples were collected from each subject. Following a 15-minute period at ambient temperature, the blood was centrifuged at 3,000 rpm for 10 minutes after coagulation had occurred for 10 minutes. After transferring the separated serum to Eppendorf containers, it was maintained at -80 °C until thawed for analysis. The concentrations of IL-6, IL-10, G-CSF, and IL-1β in serum were determined utilizing commercial ELISA sandwich kits supplied by Sunlong Biotech Co., Ltd. (Zhejiang, China). The remaining ELISA kits (IL-17, IL-21, IL-22, IL-23, and TNF-α) were obtained from Melsin Medical Co. (Jilin, China). Samples were diluted as necessary when highly concentrated biomarkers were present. The precision within the assay was indicated by a within-assay coefficient of variation of less than 10% for all assays. 1.0 pg/ml was the sensitivity of the assays for IL-10, IL-17, IL-21, IL-22, IL-23, and TNF-α; 0.1 pg/ml was the sensitivity for IL-1β and IL-6. Using these assays we constructed three different z unit composite scores: a) z score of IL-1β (z IL-1β) + z IL-6 + z TNF-α (labeled M1 macrophage function), b) z IL-17 + z IL-6 + z IL-23 (Th-17 index); and c) z IL-1β + z IL-6 + z TNF-α + z IL-17 + z IL-23 (denoted as the immune-inflammatory responses system or IRS index) [28].

### Statistics

#### Classical Statistical analysis

The researchers utilized analysis of variance (ANOVA) to examine the variations in scale variables between the three groups, namely controls and SNP and MNP. To examine the relationships between nominal variables, such as sex, and those diagnostic groups, we applied analysis of contingency tables (χ^2^-test). To ascertain the association between diagnostic classes and biomarkers and neurocognitive test results, we utilized univariate general linear model (GLM) analysis. This approach accounted for confounding variables such as nicotine dependence, sex, age, BMI, and education. The estimated marginal mean (SE) values derived by the model (GLM analysis) were calculated and presented as z scores. To distinguish the differences between biomarkers and cognitive tests between the three diagnostic groups, we conducted multiple pairwise comparisons of treatment means using the Least Significant Difference method. For statistical significance, two-tailed tests were conducted using a p-value of 0.05. All statistical analyses were conducted utilizing version 25.2017 of IBM SPSS for Windows.

### Principal component analysis (PCA)

A joint principal component analysis (PCA) was conducted on the biomarkers, cognitive tests results, and symptom domains of all participants in the current study. This involved utilizing a 20-fold cross-validation scheme, standard deviation weighting process, and singular value decomposition. The objective was to visually represent the distribution of all cases with respect to the three diagnostic classes, which are distinguished by marker color and shape. PCA was conducted using the CAMO software [36]. To represent PC scores visually, we employed various combinations of 2D and 3D dimensions (e.g., PC1 and PC2, PC1 and PC2 and PC3, PC1 and PC3, PC2 and PC3). We employ Hotelling’s T² ellipse (alpha=0.05%) in the identical 2D PC graphs to draw attention to anomalies that have the potential to impact the model. The outlier limits were established using Hotelling’s T2 and 0.05% F-residuals. The percentage of variance explained by the successively extracted PCs was then computed. In addition, several variances were examined, such as the ratio of calibrated to validated residual variance, increased limits for residual variance, and Q-residuals [36]. Subsequently, we employed the correlation loadings plot to evaluate the loadings of the input variables on the different PCs. This plot consists of two ellipses, with the exterior ellipse representing 100% of the explained variance and the inner ellipse representing 50% of the explained variance.

### Support Vector Machine (SVM) and linear discriminant analysis (LDA)

SVM is a supervised pattern recognition technique that demonstrates utility as both a learning and classification instrument. Moreover, it is frequently implemented as a data mining method. We used a 10-fold cross-validation procedure to implement SVM with a linear kernel (linear SVM) or a radial basis function kernel (RBF SVM) [36]. The model underwent validation through support vectors, which establish the most adequate demarcation between the diagnostic classes. To validate the SVM model, we also examined the outcome of the classification in a test sample (50% test and 50% training sample). The figures of merit consist of two key metrics: a) the confusion matrix, which presents the classification outcomes as predicted classes versus actual classes, and b) the classification accuracy, which is calculated as the proportion of cases correctly classified in the validation and calibration samples. We used Statistica 12.0 to run SVM. LDA is another frequently used supervised pattern recognition technique for object classification. LDA utilizes the probability distribution within the classes to generate a model or decision rule that can be implemented to assign new subjects to the most probable class. Like SVM, the confusion matrix and classification accuracy (prediction rate) are the most imperative figures of merit. Soft Independent Modeling of Class Analogy or Statistical Isolinear Multiple Component Analysis (SIMCA)

The SIMCA method is a class-modeling approach that generates confidence envelopes encircling the models of the predetermined diagnostic classes. The number of PCs utilized to construct the models is determined through cross-validation; therefore, the number of PCs may vary among the classes. Influential outliers, which are those that have a strong impact on the PCA model (e.g., when one outlier accounts for the variance in one PC while leaving most of the variance in the subsequent PC), are removed. Furthermore, subjects and influential outliers with more extreme F-residual and Hotelling’s T2 values are excluded from the analysis if their feature values are deemed irrelevant to the model. Consequently, the class PC models delineate the comparable attributes and similarities among the subjects comprising the model classes.

SIMCA computes a) model boundaries which delineate the class models constructed using PCA, and b) identification values (distances) for all cases with respect to the different class models. These values are determined by two distances: Si, or the subject to model distance, which indicates the subject’s proximity to the target class, and Hi, or the leverage of one subject to the model center, which indicates the subject’s degree of dissimilarity from the other subjects in that class. The critical distance limits for both Si and Hi are computed and employed in classification tasks through the utilization of F tests that admit members of the target class with a false negative ratio of α=0.05 (or 0.01). Thus, PCA models are then applied to patients and controls and the critical limits of Si and Hi. Subsequently, unknown or test set subjects may be projected onto these class models allowing one to authenticate cases as belonging to the target class. Cases from another class allocated to the target class are considered intruders or aliens, and cases of the target class that are not projected into the target model are considered outliers. It is possible that cases cannot be allocated to any class or are allocated to two classes. The SIMCA figures of merit consist of the following: a) the model-to-model (or intra-class) distance, which signifies the degree of dissimilarity between the models. A distance greater than three signifies that the models can be differentiated adequately, while a distance less than three indicates that the differences between the two classes are not substantial. b) The discriminatory power of the input variables is indicative of the ability of the features to distinguish between the class models, and thus the feature’s effectiveness in subject classification. This study employed two SIMCA plots: a) the Si/S0 vs Hi plot, which displays the class limits at α=0.05 and the residual standard deviation (relative distance of the subjects to the class model) versus Hi scores for a given class. b) The discriminatory power plot illustrates the extent to which the features are capable of distinguishing between the class models.

## Results

### Demographic, neurocognitive, biomarker and clinical data

Electronic Supplementary File (ESF), Table 1 shows the socio-demographic data as well as the clinical measurements in the three study groups. ESF, Table 2 shows the measurements of the neurocognitive tests (adjusted for age, sex, and education years) in the three study samples. ESF, Table 3 shows the biomarkers used in this study as well as the M1 macrophage, Th-17, and IRS indices.

### Discriminating schizophrenia patients from controls

In **Figure 1**, the PC plot resulting from the application of PCA to the three biomarker composites and eight BACS neurocognitive scores is displayed. The PC plot presents the actual subject distribution of controls and patients in a two-dimensional space, which is delineated by the first two PCs derived from the eleven variables. The PCA incorporated three biomarker indices: the index for the immune-inflammatory response system (IRS), the index for M1 macrophages, and the index for the T helper-17 axis, and the results of eight neurocognitive tests: the BACS score, digit sequencing task, list learning test, controlled word association, category instances, tower of London, symbol coding, and token motor task. PC1 and PC2 account for 83% of the variance collectively, whereas the third PC explains a mere 5% of the variance. As a result, the separation and correlation loadings of the biomarkers and neurocognitive test results on the PCs can be accurately interpreted, and this 2D diagram effectively represents the data. A substantial “street” divides the two groups: controls and patients. The controls are grouped in the left-hand section of the plot, while the patients are grouped in the right-hand section. The distribution pattern of patients and controls is comparable in the PC1-PC3 plot, which further supports the distinction between the two groups.

**Figure 1.**
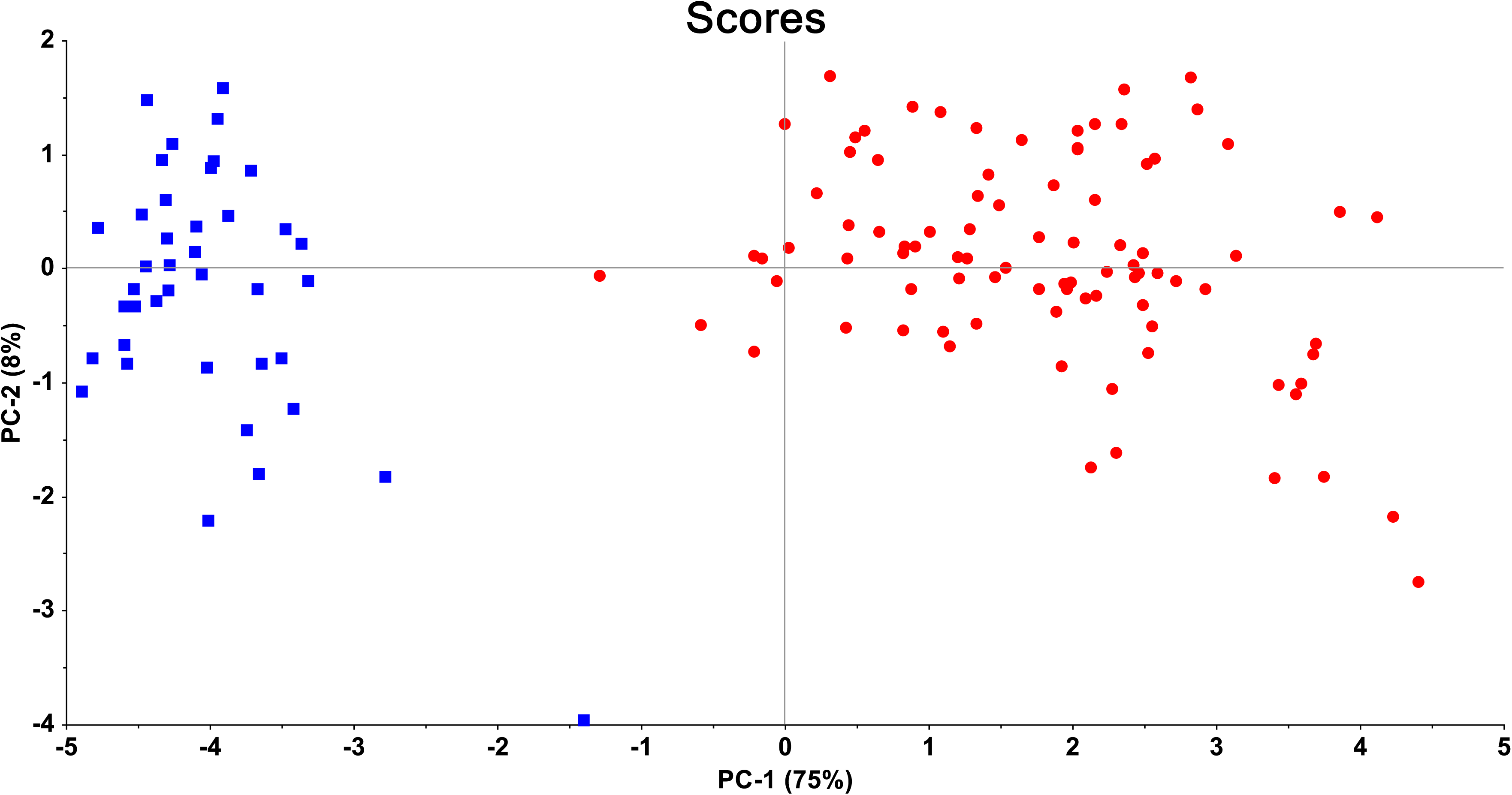
Principal Component (PC) plot obtained by PC analysis conducted on the 8 neurocognitive scores and the 3 biomarkers. This plot shows the first two PCs extracted from biomarkers and cognitive test results. Biomarkers are entered as the immune-inflammatory responses system (IRS) index, M1 macrophage index, and the T helper-17-axis index. Cognitive tests scores were the seven Brief Assessment of Cognition in Schizophrenia (BACS) tests scores as well as their total score (BACS). Class membership into schizophrenia and the normal control groups are shown by red circles and blue squares, respectively.

Nine support vectors (4 bounded) were delineated by SVM with a linear kernel: 4 vectors for schizophrenia and 5 vectors for the controls. The confusion matrix indicates that all controls and schizophrenia patients were accurately classified, indicating that the classification accuracy is 100% both prior to and following tenfold cross-validation. To assess the model’s capability to predict class membership of new subjects, the SVM analysis was reapplied to both the training (consisting of 50% patients and controls) and validation (consisting of the remaining 50%) sets. The SVM model was constructed using five support vectors, of which one was for patients and four were controls. The SVM model that was constructed demonstrated a sensitivity of 100% and a specificity of 95.7% when the validation set was projected into the training model.

SIMCA showed that the distance between the normal control model and the schizophrenia model was determined to be 146.1682, suggesting a significant distinction between the two classes. The Si/S0 versus Hi plot is illustrated in **Figure 2**. The y-axis represents the distances between the schizophrenia patients and the healthy control model, while the x-axis represents the distances from the healthy model’s center to the cases (leverage). None of the schizophrenia patients intruded upon the normal control hyperspace, and all healthy volunteers (except for one control) were authenticated as normal controls. Each of the eleven variables exhibited adequate modeling power for either the normal control or schizophrenia groups, as indicated by their respective powers exceeding 0.597. The discriminatory capacity of the eleven variables distinguishing schizophrenia from controls is illustrated in Figure 3. In descending importance, the following seven discriminatory variables were considered: the token motor task, the T helper-17 axis index, the digit sequencing task, the M1 macrophage index, controlled word association, category instances, and Tower of London.

**Figure 2.**
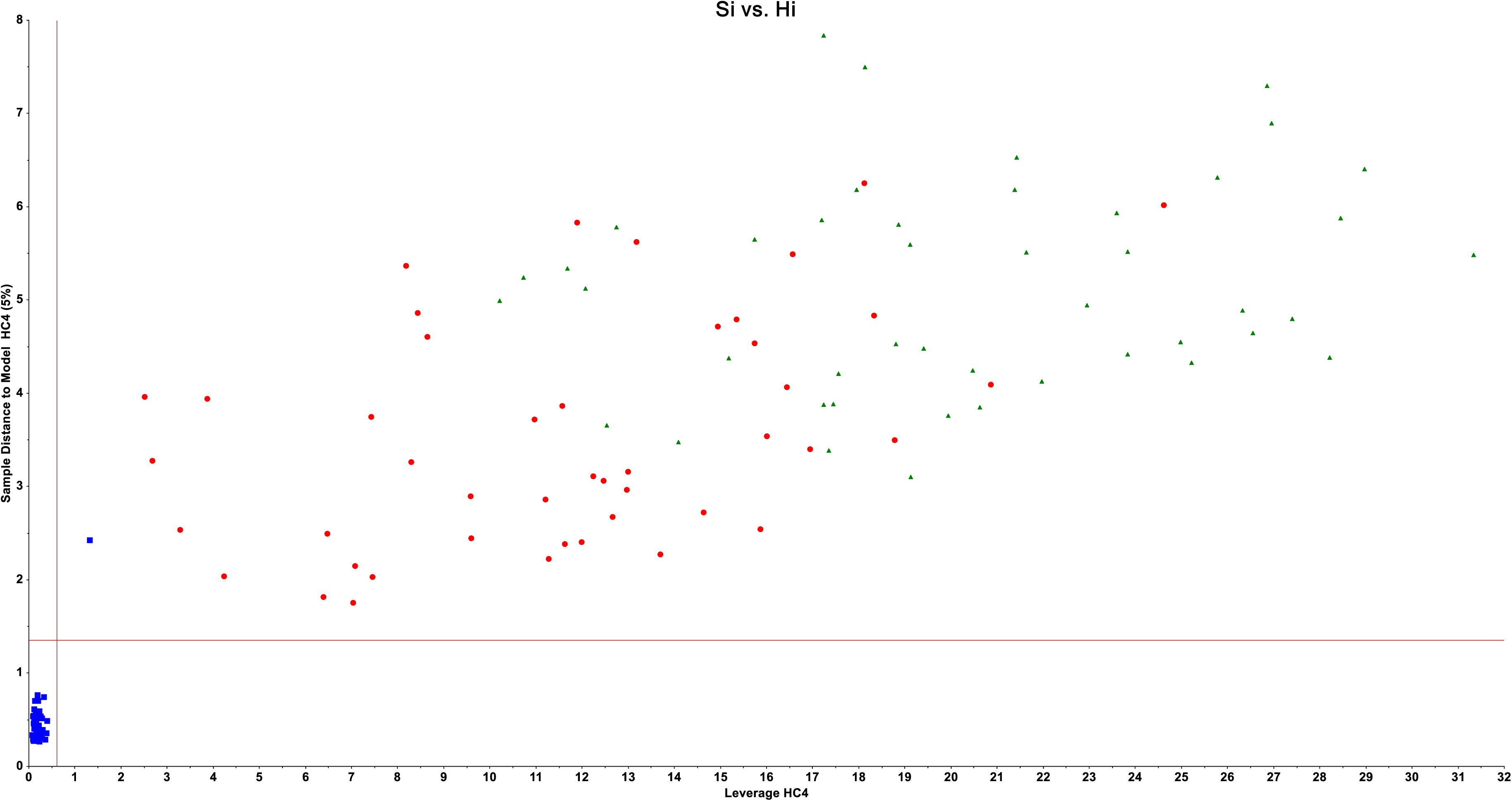
Results of Soft Independent Modelling of Class Analogy (SIMCA) displaying the Si/S0 healthy control group-membership plot (below-left quadrant depicted as blue squares). This plot shows the distances of all patients (green or red colour) to the healthy control model (y-axis) and the healthy control model centre (x axis). This plot shows that all patients were not allocated to the healthy control model, and that one healthy control was misclassified as an outlier.

**Figure 3.**
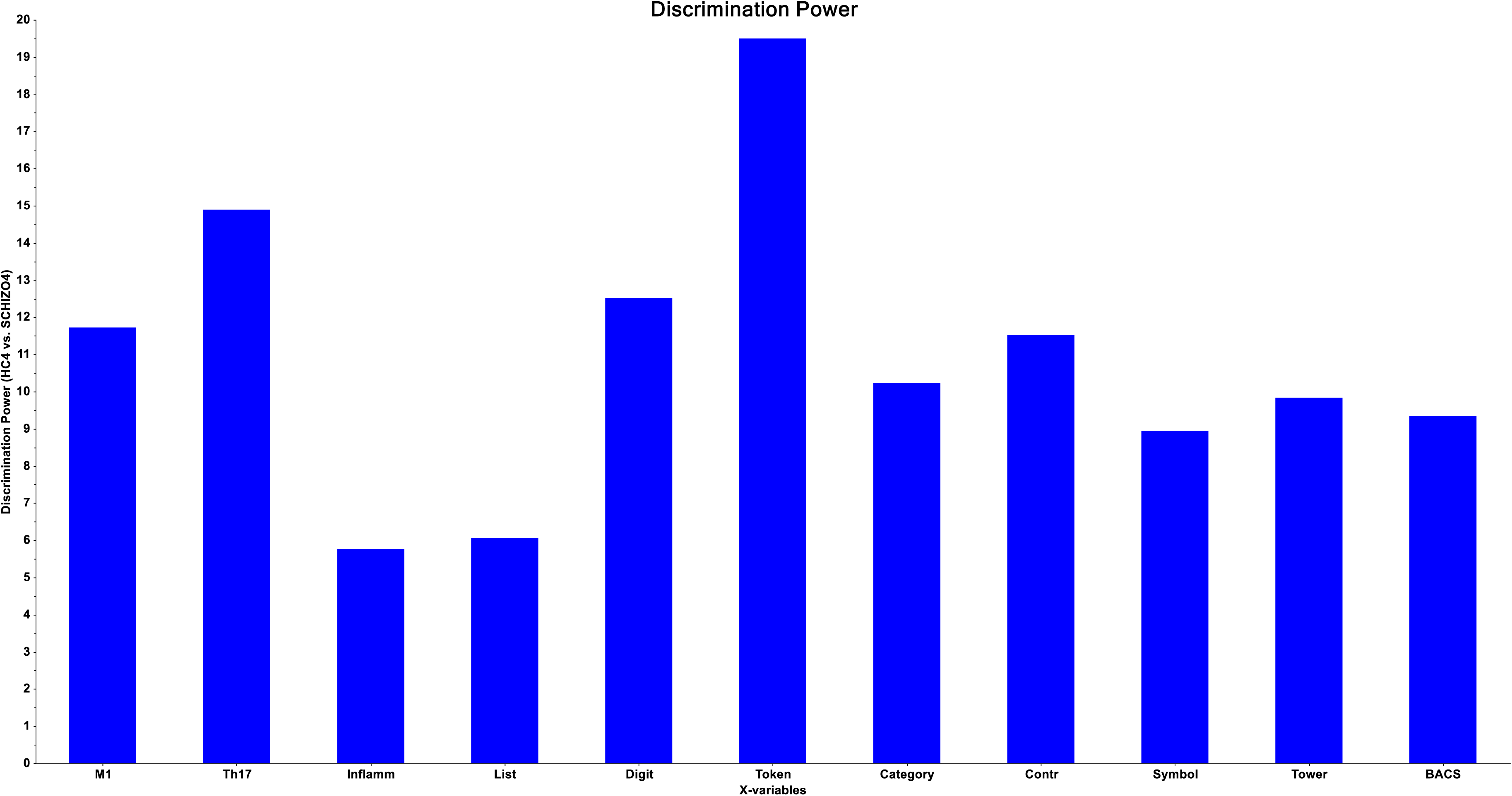
Results of Soft Independent Modelling of Class Analogy (SIMCA) showing the discrimination plot of the 3 biomarkers and 8 Brief Assessment of Cognition in Schizophrenia (BACS) test scores separating the models of schizophrenia patients versus healthy controls. Inflamm: immune-inflammatory index, M1: M1 macrophage index; Th-17: T helper-17 index, List: List learning test, Digit: Digit sequencing task, Token: Token motor task, Category: Category Instances, Contr: Controlled word association, Symbol: Symbol coding, Tower: Tower of London, BACS: total composite score on all 7 BACS items.

### Discrimination of MNP, SNP and controls using biomarkers and cognitive tests

**Figure 4** presents an identical PC plot as Figure 1, with the exception that it differentiates between control and MNP and SNP samples (instead of controls versus schizophrenia). The variables utilized in constructing the PC plot in Figure 1 (eight neurocognitive scores and three biomarkers) were precisely the same as those utilized in this PC analysis. The plot illustrates that MNP patients formed a cluster based on the three biomarkers and eight cognitive test results and assembled at the right side of the diagram. Similarly, SNP patients exhibited a clustering pattern. However, a distinct demarcation between the two categories was absent. SVM demonstrated that MNP and SNP could be distinguished by employing 38 vectors (20 for SNP and 18 for MNP). The classification accuracy of this 10-fold cross-validated SVM model was 91.11% for SNP patients and 97.78% for MNP patients. LDA performed on the three study groups and using the 11 input variables yielded an accuracy of 96.92%. All controls were correctly classified, 91.1% of the SNP patients, and 95.6% of the MNP patients. Electronic supplementary File (ESF) Figure 1 shows the outcome of this LDA.

**Figure 4.**
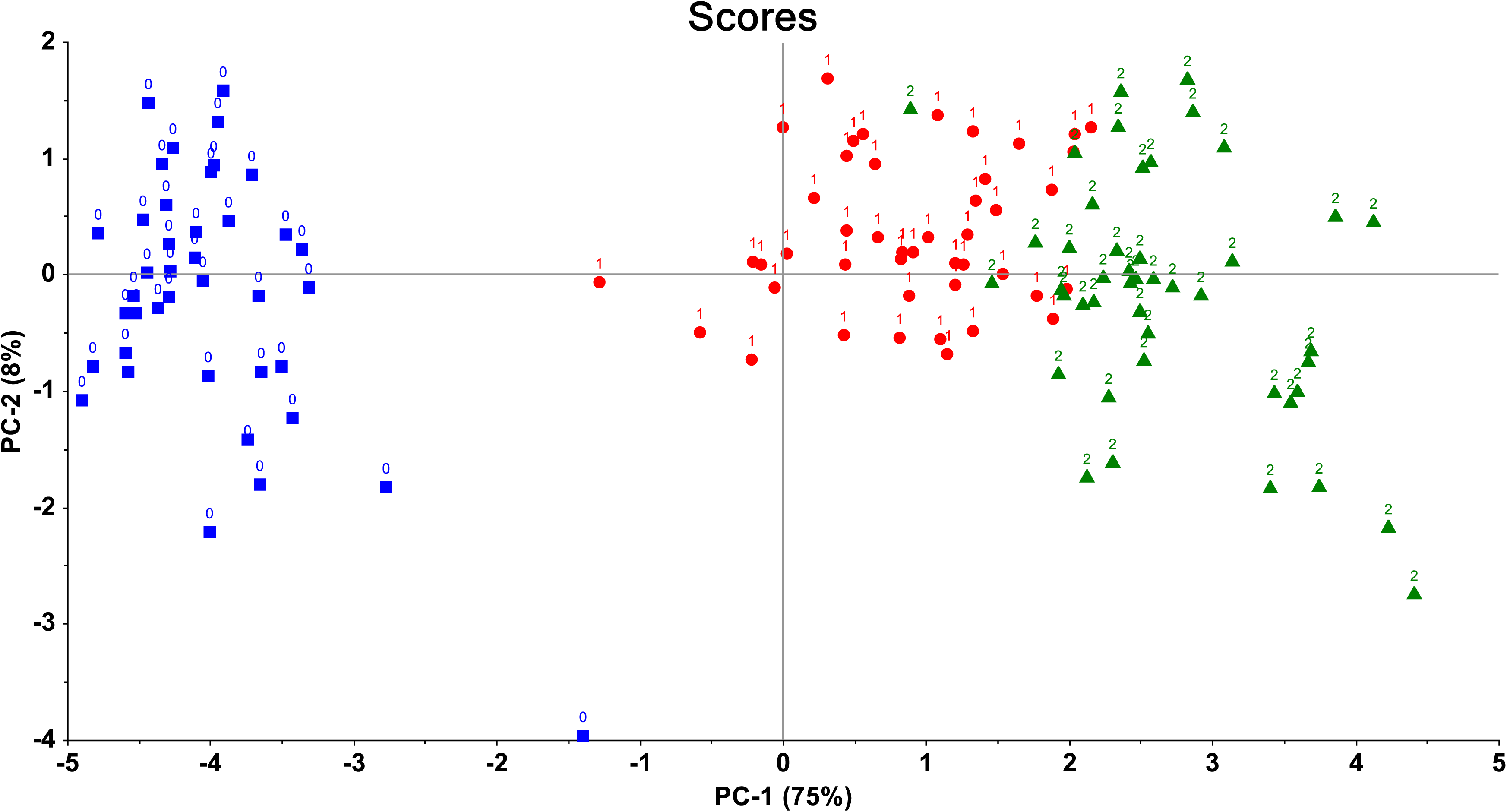
The Principal Component (PC) plot shown in Figure 1 but now displaying the distribution of patients with major neurocognitive psychosis (MNP) and simple psychosis (SP) versus healthy controls. This PC analysis was conducted on the 8 neurocognitive scores and the 3 biomarkers. This plot shows the first two PCs extracted from 3 biomarkers (immune-inflammatory responses system) index, M1 macrophage index, and the T helper-17-axis index. Cognitive tests scores were the seven Brief Assessment of Cognition in Schizophrenia (BACS) tests scores as well as their total score. Normal control groups: blue squares; red circles: SP; and green triangles: MNP.

The demarcation of MNP from SNP and controls is further substantiated when one looks at the three-dimensional plots showing the distribution of all cases with respect to the three classes (see **Figure 5 and ESF, Figure 2**). These figures are three-dimensional plots with PC1, PC2, PC3 or PC1, PC3, PC4 scores of all subjects. It can be observed that all controls cluster together and that the distribution of MNP is quite different from that of SNP.

**Figure 5.**
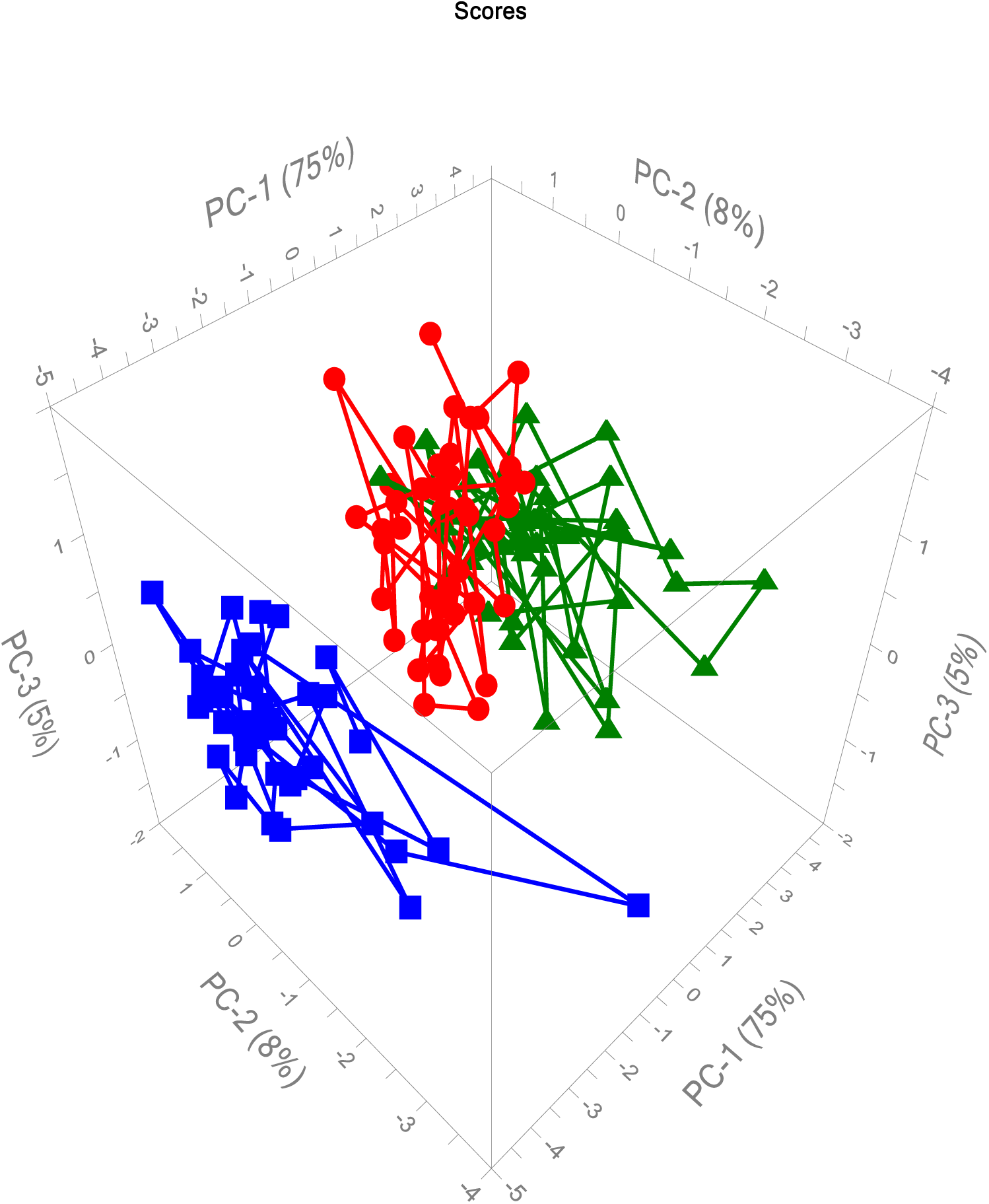
Three-dimensional plot showing the distribution of all normal controls (blue squares), patients with simple neurocognitive psychosis (SNP, red circles) and major neurocognitive psychosis (green triangles) with respect to principal component 1 (PC1), PC2, and PC3, which were extracted from three biomarkers and 8 cognitive tests scores.

SIMCA showed that the distance between the MNP and SNP model was 16.1269, indicating a significant distance between both models and, therefore, significant distinctions between the two classes. The discriminatory capacity of the eleven variables distinguishing MNP from SNP is illustrated in **Figure 6**. In descending order of importance, the following discriminatory variables were considered: M1 macrophage profile, category instances, Tower of London, symbol coding, and list learning task. The presented data indicates that the MNP and SNP classes represent distinct patient groups in terms of biomarkers and cognitive functions.

**Figure 6.**
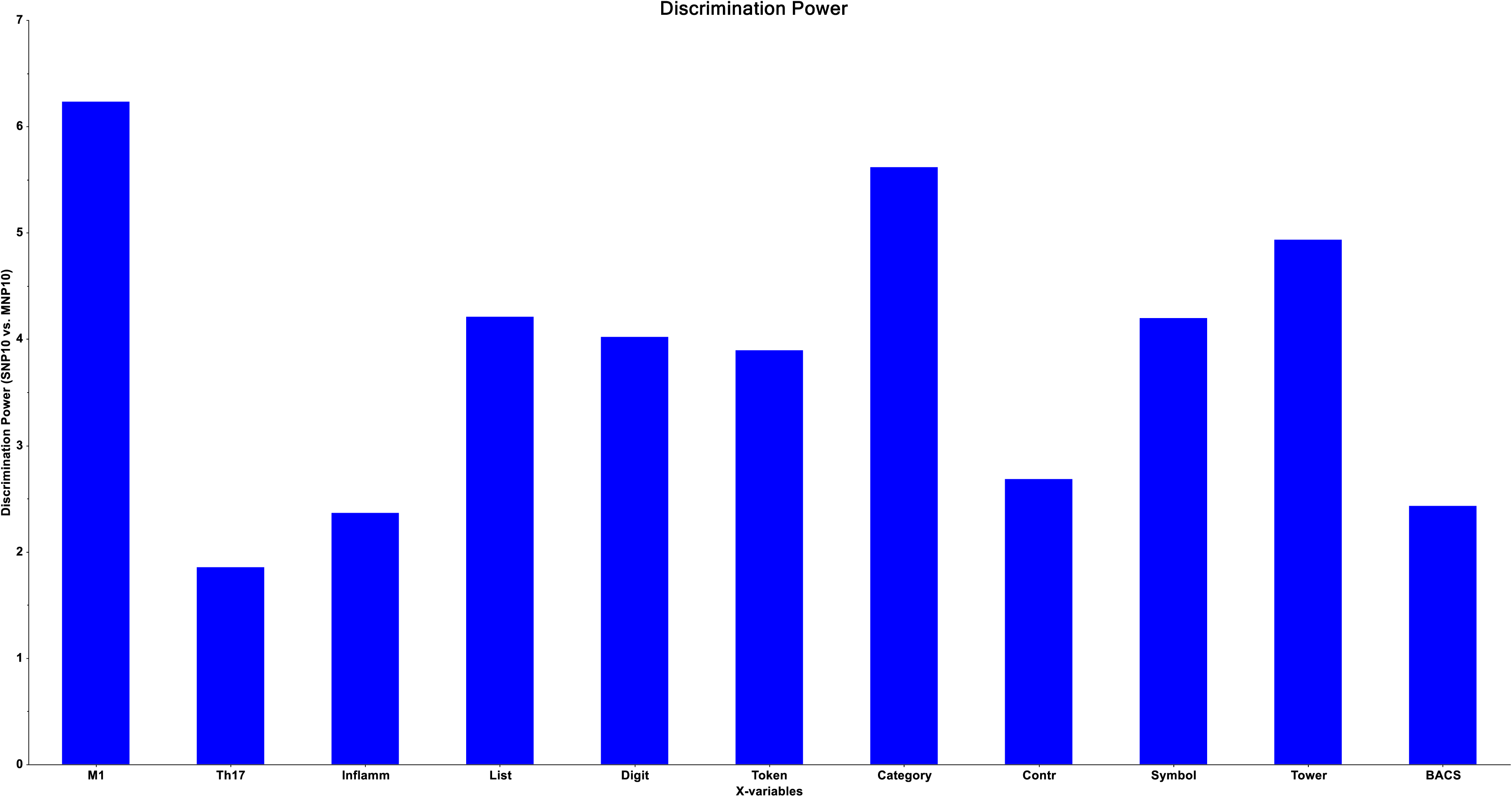
Results of Soft Independent Modelling of Class Analogy (SIMCA) showing the discrimination plot of the 3 biomarkers and 8 Brief Assessment of Cognition in Schizophrenia (BACS) test scores separating the models of major neurocognitive psychosis from simple neurocognitive psychosis. Inflamm: immune-inflammatory index, M1: M1 macrophage index; Th-17: T helper-17 index, List: List learning test, Digit: Digit sequencing task, Token: Token motor task, Category: Category Instances, Contr: Controlled word association, Symbol: Symbol coding, Tower: Tower of London, BACS: total composite score on all 7 BACS items.

### Discrimination of MNP, SNP and controls using biomarkers, cognitive and clinical subdomains

The PC plot presented in **Figure 7** is the result of a PC analysis performed on eighteen input variables, including seven clinical domains, and the three biomarkers, and eight neurocognitive scores as described above. Considering that both PCs accounted for 82% of the variance in the data set, this 2D plot accurately represents the data. As previously mentioned, patients and controls are effectively segregated, and similarly, MNP and SNP patients are divided into two distinct clusters. Four MNP patients, however, were incorrectly classified as SNP patients. The correlation loading diagram, as illustrated in **Figure 8**, presents the correlation loadings among the 18 input variables on the first two PCs. The correlation between PC1 and the biomarkers and symptom subdomains were positive, whereas the association between PC1 and the neurocognitive symptoms was inverse. Therefore, differentiation between patients with schizophrenia and controls, as well as between patients with MNP and SNP, is mainly achieved by utilizing elevated scores on biomarkers and symptom subdomains, and decreased scores on cognitive tests.

**Figure 7.**
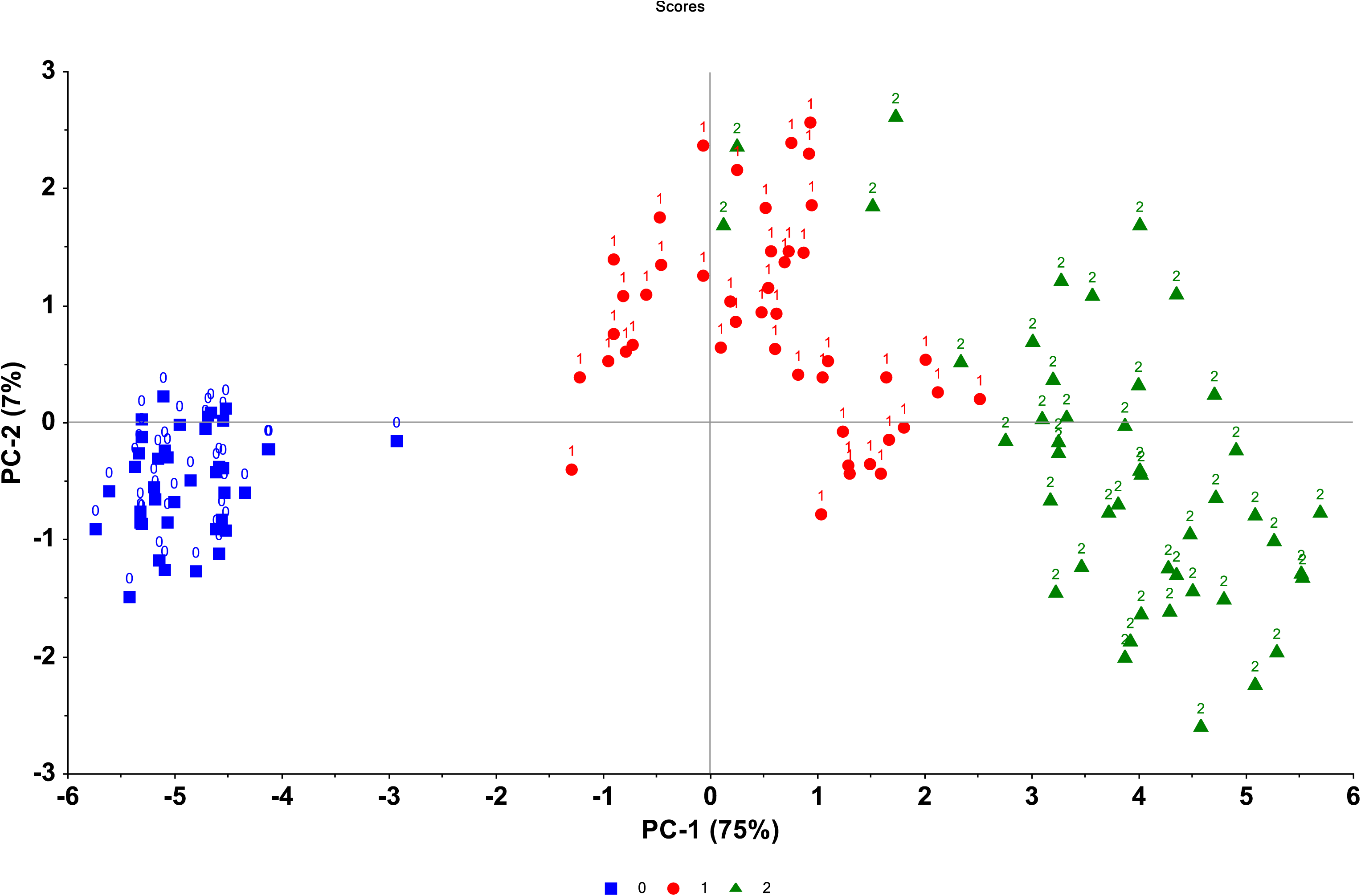
Principal Component (PC) plot obtained by PC analysis conducted on the 8 neurocognitive scores, 3 biomarkers, and 7 clinical domains. This figure shows the distribution of patients with major neurocognitive psychosis (MNP) and simple psychosis (SP) versus healthy controls. This plot shows the first two PCs extracted from biomarkers, cognitive test results and clinical subdomains. Biomarkers are entered as the immune-inflammatory responses system index, M1 macrophage index, and the T helper-17-axis index. Cognitive tests scores were the seven Brief Assessment of Cognition in Schizophrenia (BACS) as well as their total composite score (BACS). The clinical subdomains comprised psychosis, hostility, excitation, mannerism, negative symptoms, formal thought disorders, and psychomotor retardation. Normal controls are shown as blue squares; SP: as red circles: MNP as green triangles.

**Figure 8.**
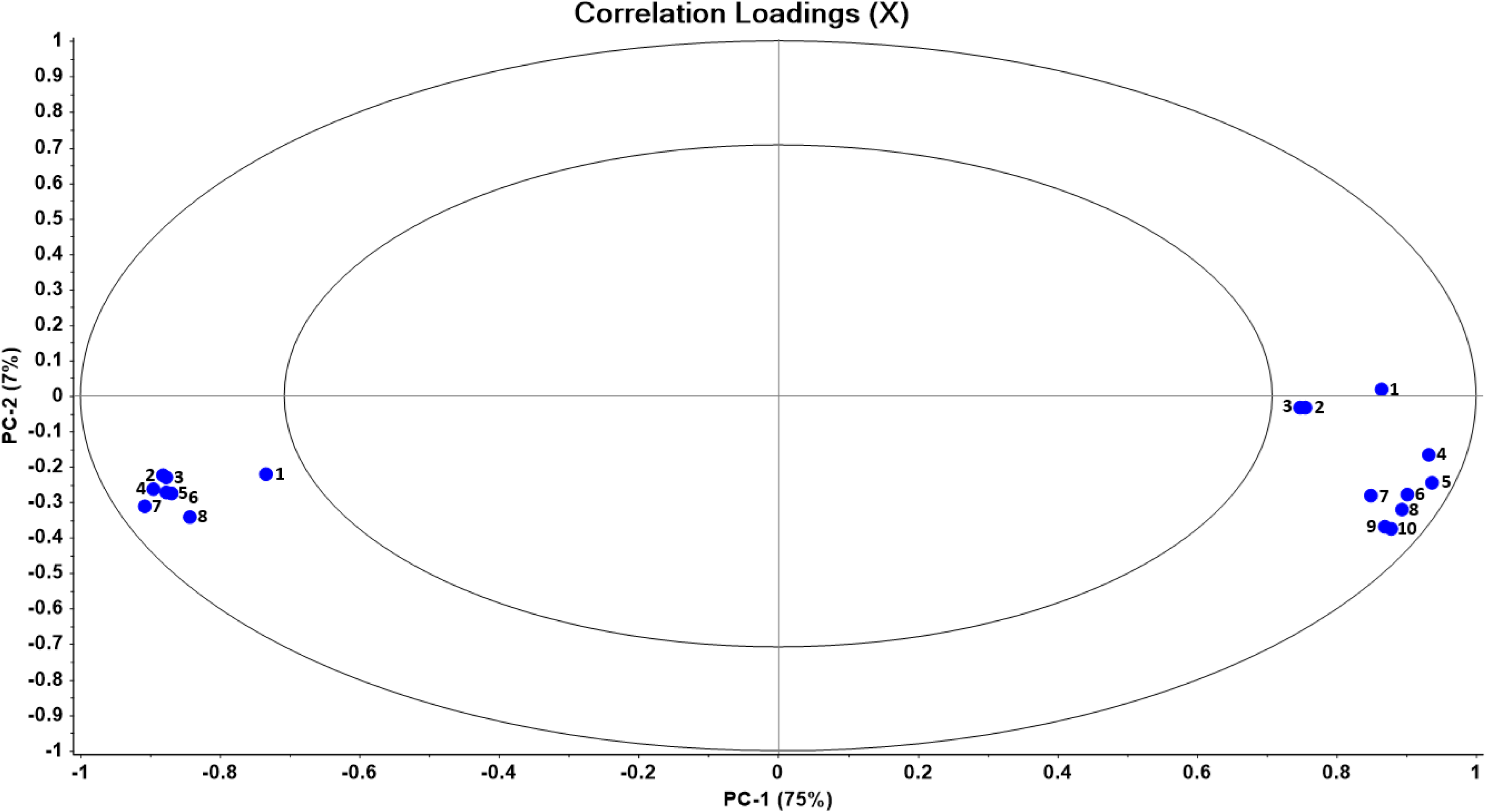
Correlation loading plot. This plot shows the correlation loadings of 18 variables on the first two principal components (PC) displayed in Figure 5. The 18 variables, which were used to construct the PC model shown in Figure 5, comprise: 8 neurocognitive scores, 3 biomarkers, and 7 clinical domains. This plot displays two ellipses, the outer one indicating 100% explained variance, and the inner ellipse indicating 50% of the explained variance. The 10 variables shown on the right site of the plot are: 1: immune-inflammatory index, 2: M1 macrophage index; 3: T helper (Th)-17 index, 4: negative symptoms; 5: psychosis; 6: psychomotor retardation, 7: excitation, 8: formal thought disorders, 9: mannerism; and 10: hostility. The 8 variables shown on the left site of the plot are the Brief Assessment of Cognition in Schizophrenia (BACS) probes: 1: Controlled word association, 2: Symbol coding, 3: Token motor task, 4: Tower of London test, 5: List learning test, 6: Digit sequencing task, 7: Total composite score, 8: Category Instances.

SVM (linear kernel with 10-fold cross-validation) demonstrated that using 36 support vectors, all controls, and SNP patients were accurately classified, while 95.6% of all MNP patients were correctly classified. LDA revealed that 96.92% of all cases, including all controls, 95.6% of all SNP patients, and 95.6% of MNP patients, were correctly classified.

The outcome of SIMCA using the Si/S0 versus Hi model of MNP is displayed in **Figure 9**, specifically in the lower-left quadrant with green triangles representing MNP patients. The distances between SNP patients and controls to the MNP model and leverage are depicted in the y- and x-axis, respectively. None of the SNP patients or controls intruded upon the MNP model, as shown in this graph. However, it was not possible to authenticate three MNP patients as members of the MNP class; therefore, these patients should be considered outliers and, thus, misclassifications. In the lower-left quadrant of **Figure 10**, the Si/S0 versus Hi model of the SNP class is illustrated, with SNP patients denoted by red circles. Four MNP patients were misclassified as aliens because of their intrusion into the SNP model. In the SNP model, none of the typical controls were allocated. The distance between the MNP and SNP models was 7.117, which is a statistically significant distance indicating separation of the study groups. In descending order of importance, the five most discriminatory input variables were as follows: token motor task, mannerism, Th-17 axis, Tower of London, and excitation. **Figure 11** is a three-dimensional plot which shows the PC scores of all cases on PC1, PC2, PC3. It can be observed that the SNP and MNP cluster together and show a distinct distribution of the data points into this three-dimensional space. We rerun another SIMCA using the 4 most variables discriminating the groups as modelling and discriminating variables, namely token motor task, mannerism, Th-17 axis, and Tower of London. We found that the MNP model to SNP model distance was 14.66.

**Figure 9.**
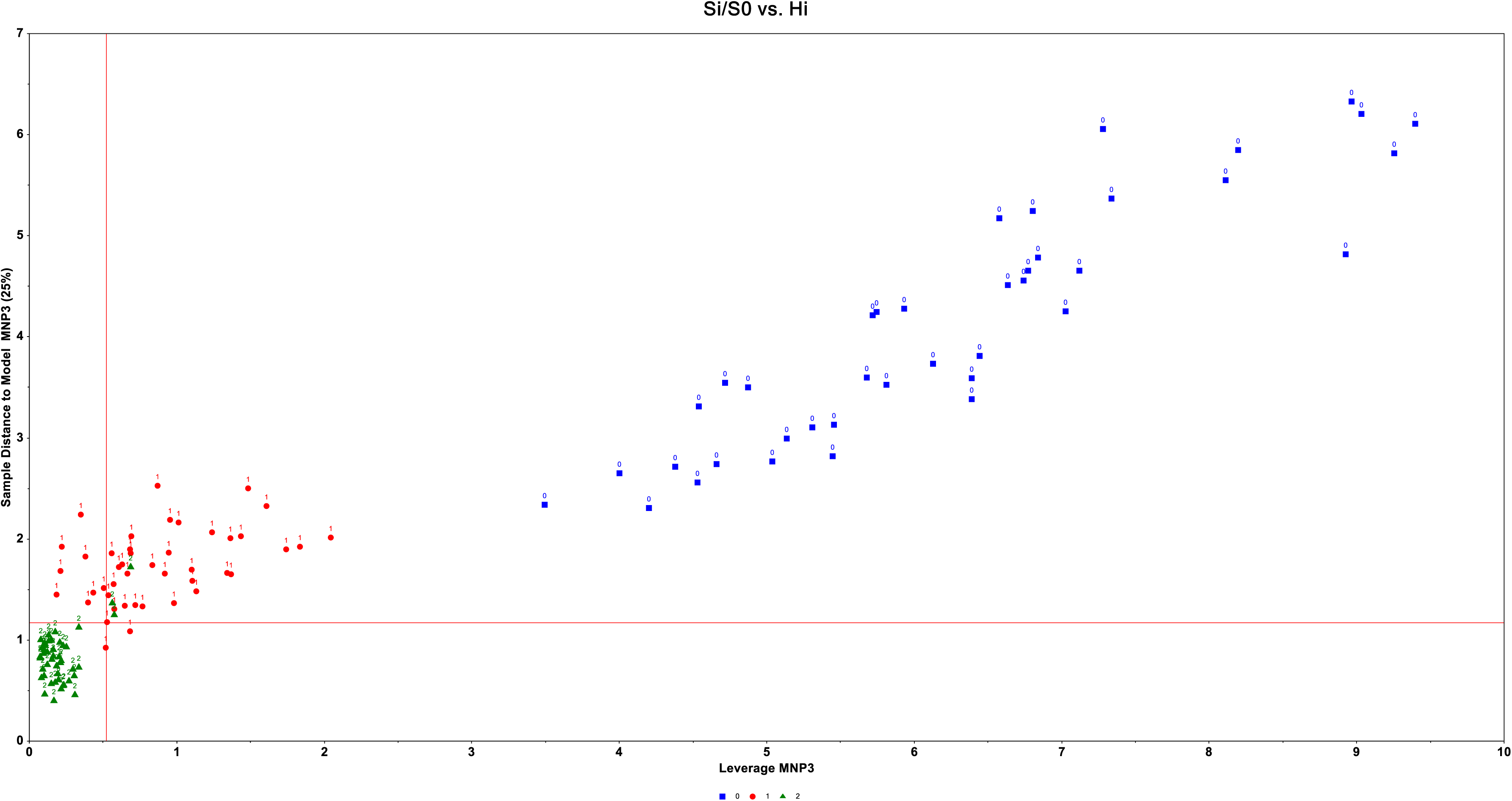
Results of Soft Independent Modelling of Class Analogy (SIMCA) displaying the Si/S0 simple neurocognitive psychosis (SNP) model (below-left quadrant depicted as red circles). This plot shows the distances of major neurocognitive psychosis (MNP, green triangles) and healthy controls (blue squares) to the SNP model, constructed using SIMCA (scores on the y-axis) and the SNP leverage or centre (scores on the x axis). This plot shows that three MNP patients could not be authenticated as MNP patients, whilst all SNP patients and controls were not allocated to the MNP model.

**Figure 10.**
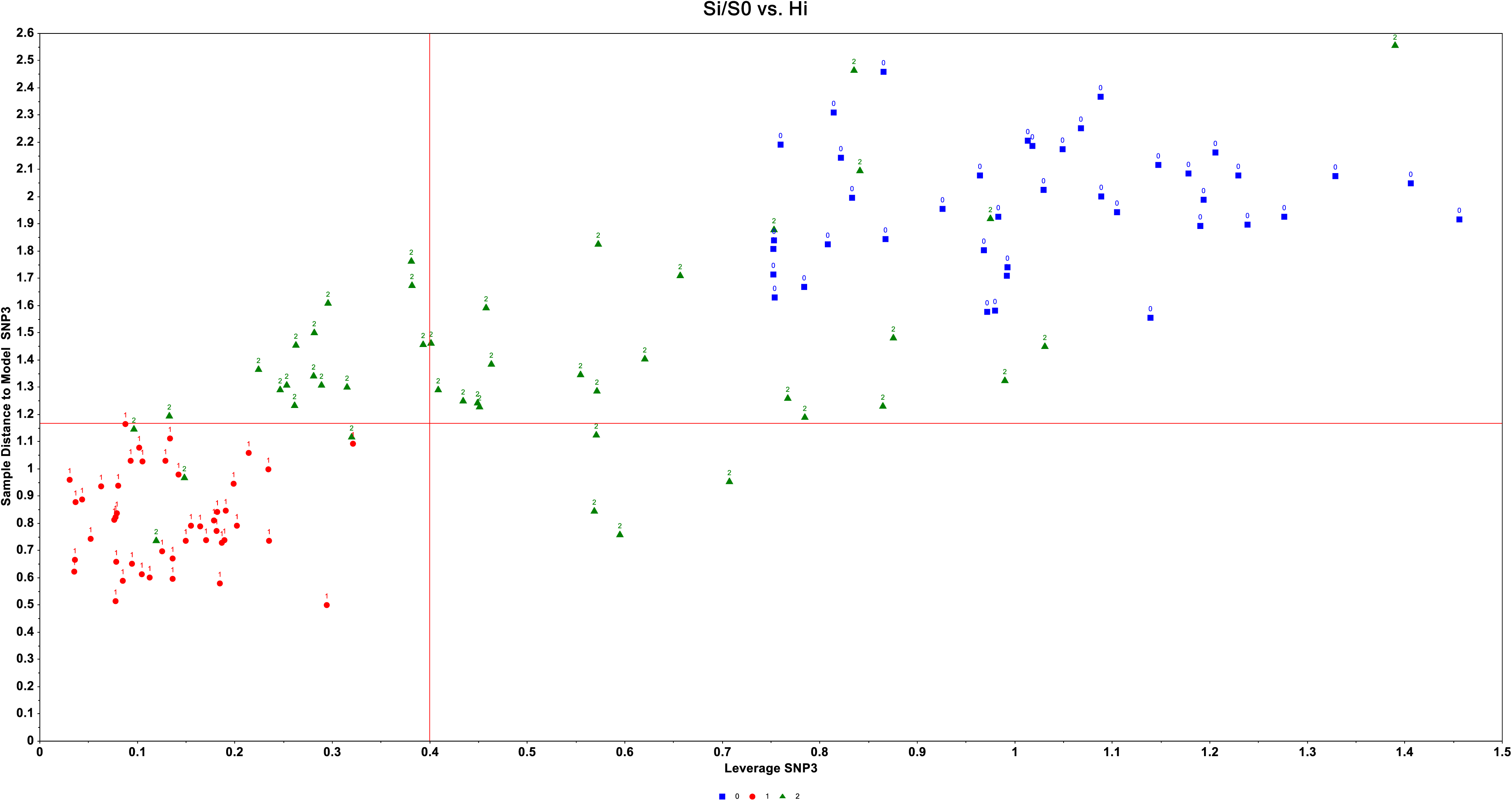
Results of Soft Independent Modelling of Class Analogy (SIMCA) displaying the Si/S0 major neurocognitive psychosis (MNP) model (below-left quadrant depicted as green triangles). This plot shows the distances of simple neurocognitive psychosis (SNP, red circles) and healthy controls (blue squares) to the MNP model, constructed using SIMCA (scores on the y-axis) and the MNP leverage or centre (scores on the x axis). This plot shows that all SNP patients could be authenticated as SNP patients (there were no outliers), whilst four MNP patients were allocated to the SNP model, and thus were misclassifications.

**Figure 11.**
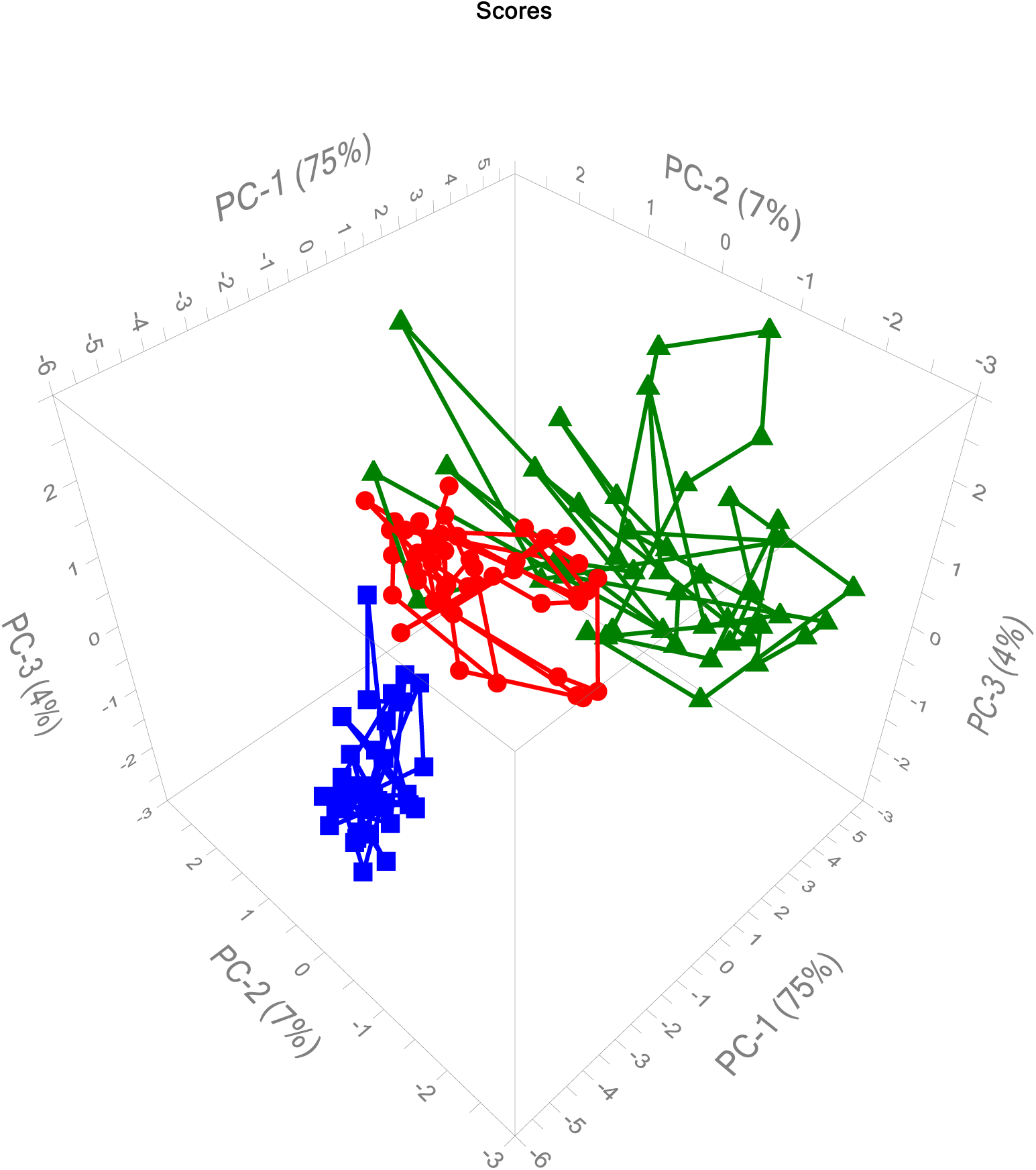
Three-dimensional plot showing the distribution of all normal controls (blue squares), patients with simple neurocognitive psychosis (SNP, red circles) and major neurocognitive psychosis (green triangles) with respect to principal component 1 (PC1), PC2 and PC3, which were extracted from three biomarkers, 8 cognitive tests scores, and 7 symptom subdomains.

## Discussion

### MNP and SNP are qualitative distinct categories

Based on neurocognitive assessments and the IL-6/IL-23/Th-17 axis, we found that MNP and SNP are qualitatively distinct classes. Our analysis reveals that the class models for SNP and MNP are distributed in distinct subspaces within the multivariate space generated by the neurocognitive and biomarker assessment data sets. In addition, SIMCA showed that the distances between the models constructed around patients with psychosis and controls were statistically significant.

Previous research has also shown that MNP and SNP are classified differently when it comes to neurotoxic TRYCATs and neurocognitive assessments [37]. In the present study, the immune-associated neurotoxicity is increased in MNP as compared with SNP and controls, as indicated by the IL-6/IL-23/Th-17 axis [25, 38]. Therefore, it appears that MNP is characterized by immune-associated neurotoxicity with regard to the Th-17 and TRYCAT pathways [21]. In a study conducted by Kanchanatwan et al., it was found that the difference between the SNP and MNP models, which were based on biomarkers (TRYCAT levels) and neurocognitive deficits, was only moderate at 4.59 [37]. In the current study, the model-to-model distance between both MNP and SNP was found to be 16.13, which is significantly higher. Based on this finding, the Th-17 axis is probably more prominent in the MNP features than the TRYCAT pathway. In a study conducted by Al-Hakeim et al. (2020), the plasma levels of various cytokines and chemokines were analyzed in 120 individuals with MNP and 54 healthy controls [39]. The researchers also examined the relationship between these levels and neurocognitive scores, as measured by the Brief Assessment of Cognition in Schizophrenia, as well as PHEMN symptoms. The study successfully differentiated the MNP group from the healthy control group based on these factors yielding a SIMCA distance of 19.3 between the models that represented the healthy control class and the MNP class. This indicates a significant distinction between the models constructed for these two groups. However, in the present study, a notable distance of 146.1682 was observed between the groups diagnosed with schizophrenia and the control group. Therefore, the IL-6/IL-23/Th-17 axis may have a greater impact on MNP compared to the approach used in the study conducted by Al-Hakeim et al. (2020), which focused on the combination of neurotoxic M1 cytokines with CC2 and CCL11.

### Why using PCA and SIMCA

In previous studies, Maes et al. have highlighted the utility of classical statistical tests such as ANOVA and GLM analysis in detecting quantitative differences in biomarkers among groups. However, it is important to note that this approach is based on false premises, as pointed out by Maes and Anderson and Maes [21, 22]. This approach involves using the outcome data (the phenome of schizophrenia) as an explanatory variable, while considering the causal pathways that predict or delineate the phenome as the dependent variables. Machine learning techniques like binary logistic regression analysis, SVM, neural networks, and discriminant analysis are better suited for predicting the phenome of the disorder using AOP (adverse outcome pathway) variables [21, 39]. In addition, these methods can be utilized to categorize patients and controls and calculate the corresponding accuracy of the prediction by using the confusion matrix, either obtained by cross-validation or in a holdout data set. However, ML models are not suited for analyzing the actual distances between the class models surrounding the patients or controls [28].

As previously mentioned, the recommended approach is to utilize a combination of joint PCA plot (or similar ML methods) and SIMCA analyses to evaluate the distinction between classes [27, 28]. Firstly, PCA plot enables the visual evaluation of the differences in the distribution of the classes in the multidimensional space. Thus, the significant gaps between classes, such as those between controls and MNP/SNP, and between MNP and SNP, indicate that these groups can be considered as separate classes. Similarly, variations in the arrangement of the classes in various spaces (such as the PC1, PC2, and PC3 space, and the PC1, PC2, and PC4 space) indicate clear-cut qualitative classes with no or minimal overlap. Additionally, SIMCA holds great significance as it serves as a class modeling technique that constructs PCA models of the groups by conducting PCA on all pertinent features, including neurocognitive test scores and biomarkers. Consequently, the SIMCA classes are defined by PCA models in a hyperspace, and the computation of specific critical limits enables the classification of subjects and the calculation of interclass distances, which provide insights into the differences between the models. Therefore, a model-to-model distance greater than three suggests that the models can be sufficiently differentiated.

One of the benefits of SIMCA is the ability to calculate the discriminatory power of the variables that distinguish the models between the groups. Our findings indicate that there are three key markers that differentiate the MNP/SNP groups from the controls: lowered motor speed, activated Th-17, and lowered working memory. Additionally, we observed that M1 activation, lowered verbal fluency, and executive functions are the prominent features of MNP compared to SNP. However, SIMCA is considered to be less effective for classification purposes when compared to SVM and neural networks [28].

### Clinical significance

There are notable similarities between the MNP class, and the concepts of dementia praecox and deficit models as proposed by Kraepelin, Bleuler, and Snezhnevsky (see Introduction). These authors observed that a portion of patients with psychosis experience a deficit caused by psycho-organic pathophysiology. Furthermore, this study, along with previous SIMCA studies [37, 39], has characterized MNP as an immune-linked neurotoxic disease with the “defect” or “deficit” being of neuro-immune origin.

Furthermore, the decision to combine the MNP and SNP case definitions into a single entity in the DSM-IV to DSM-5 leads to inaccurate conclusions, as pointed out by Maes and Anderson (2012) [22] and Maes (2023) [21]. As an illustration, when attempting to apply a pathway that is specific to MNP or SNP to the combined group, it is possible for inaccurate results to manifest as false positives. There is a possibility of missing actual aberrations in MNP or SNP when these classes are combined, leading to false negatives [22]. The sheer magnitude of inaccurate findings in schizophrenia research, particularly in genetic and biomarker studies, which have been published due to the oversight of the MNP and SNP classes is probably highly significant. Therefore, it is crucial to re-evaluate the genetic and biomarker research conducted in schizophrenia using the MNP and SNP case definitions.

It is worth noting that the diagnostic label “schizophrenia” carries a significant stigma and fails to fully encompass the key characteristics of the disease, such as the presence of neurocognitive deficits and psychosis. Therefore, using the term “neurocognitive psychosis” is more objective and highlights the specific features of the disorder. To enhance diagnostic precision, it is advisable for clinicians to utilize the terms MNP and SNP instead of schizophrenia. This distinction allows for a more accurate classification, as MNP is associated with greater neurocognitive deficits, heightened severity of psychosis, and more profound pathway abnormalities. Therefore, it is advisable to utilize our machine learning models (specifically SVM) for the purpose of categorizing patients as MNP or SNP.

## Limitations

Based on the findings of this study, it is evident that the distinction between MNP and SNP is quite pronounced. However, it is worth noting that incorporating additional characteristics of MNP could potentially enhance the accuracy of differentiating between MNP and SNP. These factors encompass a range of immune functions, such as the compensatory immunoregulatory system (CIRS), innate immune defense dysfunctions, oxidative stress, antioxidant levels, the complement system, bacterial translocation, and reduced neuroprotection [21]. In addition, future ML analyses should incorporate functional magnetic resonance imaging data, specifically focusing on abnormalities in the connectome, such as the central executive network, default mode network, and the salience network [40].

## Supporting information

Electronic Supplementary File

## Data Availability

The dataset generated during and/or analyzed during the current study will be available from the corresponding author upon reasonable request and once the dataset has been fully exploited by the authors.

## Acknowledgments

We acknowledge the staff of Al-Hakeem General Hospital, Psychiatric Unit in Najaf Governorate, for their help in collecting samples. We also acknowledge the work of the highly skilled staff of Asia Clinical Laboratory in Najaf city for their help in the ELISA assays.

## Author**’**s contributions

All the contributing authors have participated in the preparation of the manuscript.

## Research involving human participants!

Approval for the study was obtained from the Institutional Review Board of the University of Kufa, Iraq (82/2020)

## Informed consent

All participants and first-degree relatives of participants with schizophrenia gave written informed consent before participation in our study.

## Data Access Statement

The dataset generated during and/or analyzed during the current study will be available from the corresponding author (MM) upon reasonable request and once the dataset has been fully exploited by the authors.

