## Supplementary material for "The novel pathway phenotype “major neurocognitive psychosis” is validated as a distinct class through the analysis of immune-linked neurotoxicity biomarkers and neurocognitive deficits": Electronic Supplementary File: ESF-Major neurocognitive psychosis.pptx

### Slide 1
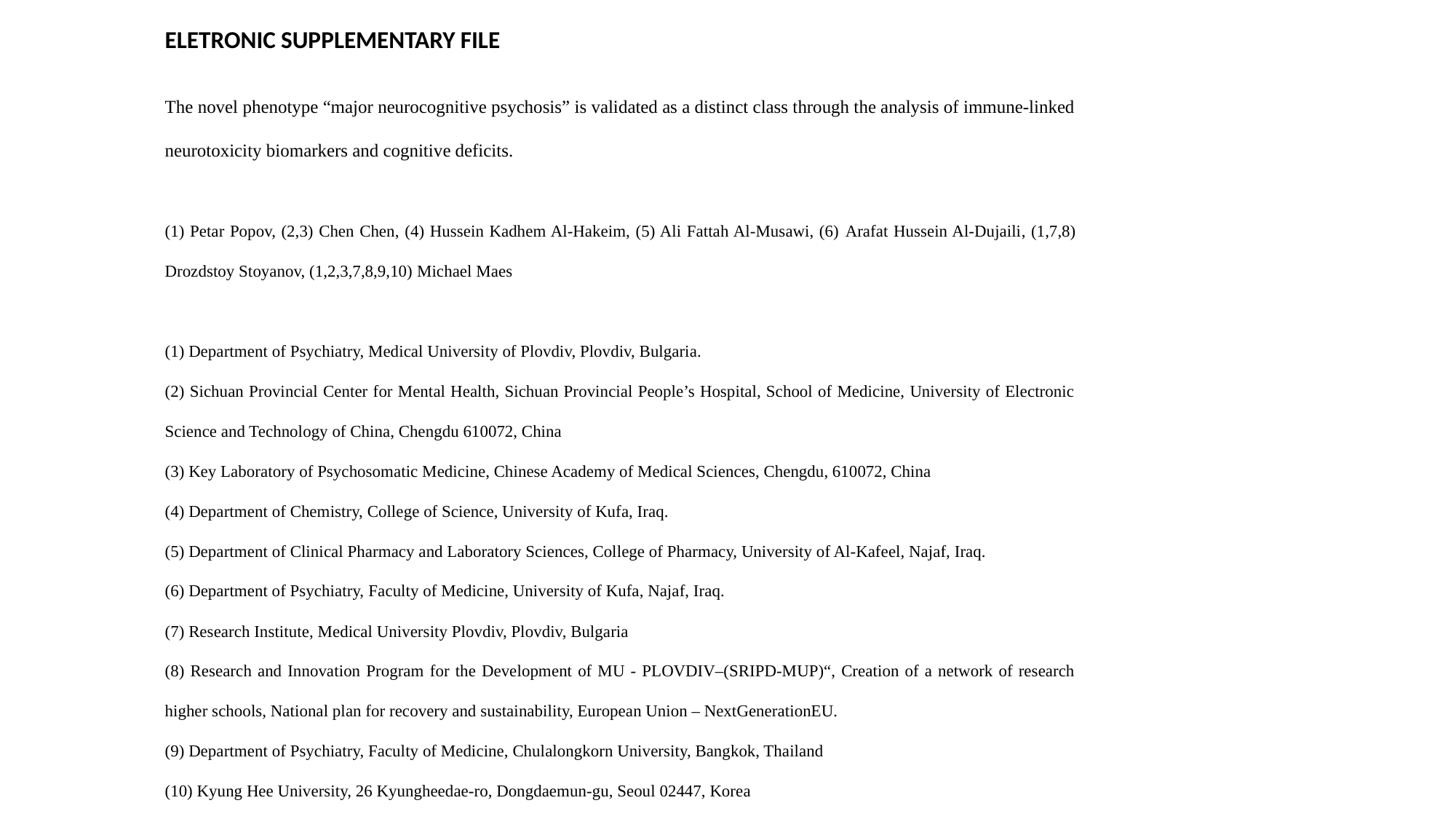

ELETRONIC SUPPLEMENTARY FILE
The novel phenotype “major neurocognitive psychosis” is validated as a distinct class through the analysis of immune-linked neurotoxicity biomarkers and cognitive deficits.
(1) Petar Popov, (2,3) Chen Chen, (4) Hussein Kadhem Al-Hakeim, (5) Ali Fattah Al-Musawi, (6) Arafat Hussein Al-Dujaili, (1,7,8) Drozdstoy Stoyanov, (1,2,3,7,8,9,10) Michael Maes
(1) Department of Psychiatry, Medical University of Plovdiv, Plovdiv, Bulgaria.
(2) Sichuan Provincial Center for Mental Health, Sichuan Provincial People’s Hospital, School of Medicine, University of Electronic Science and Technology of China, Chengdu 610072, China
(3) Key Laboratory of Psychosomatic Medicine, Chinese Academy of Medical Sciences, Chengdu, 610072, China
(4) Department of Chemistry, College of Science, University of Kufa, Iraq.
(5) Department of Clinical Pharmacy and Laboratory Sciences, College of Pharmacy, University of Al-Kafeel, Najaf, Iraq.
(6) Department of Psychiatry, Faculty of Medicine, University of Kufa, Najaf, Iraq.
(7) Research Institute, Medical University Plovdiv, Plovdiv, Bulgaria
(8) Research and Innovation Program for the Development of MU - PLOVDIV–(SRIPD-MUP)“, Creation of a network of research higher schools, National plan for recovery and sustainability, European Union – NextGenerationEU.
(9) Department of Psychiatry, Faculty of Medicine, Chulalongkorn University, Bangkok, Thailand
(10) Kyung Hee University, 26 Kyungheedae-ro, Dongdaemun-gu, Seoul 02447, Korea

### Slide 2
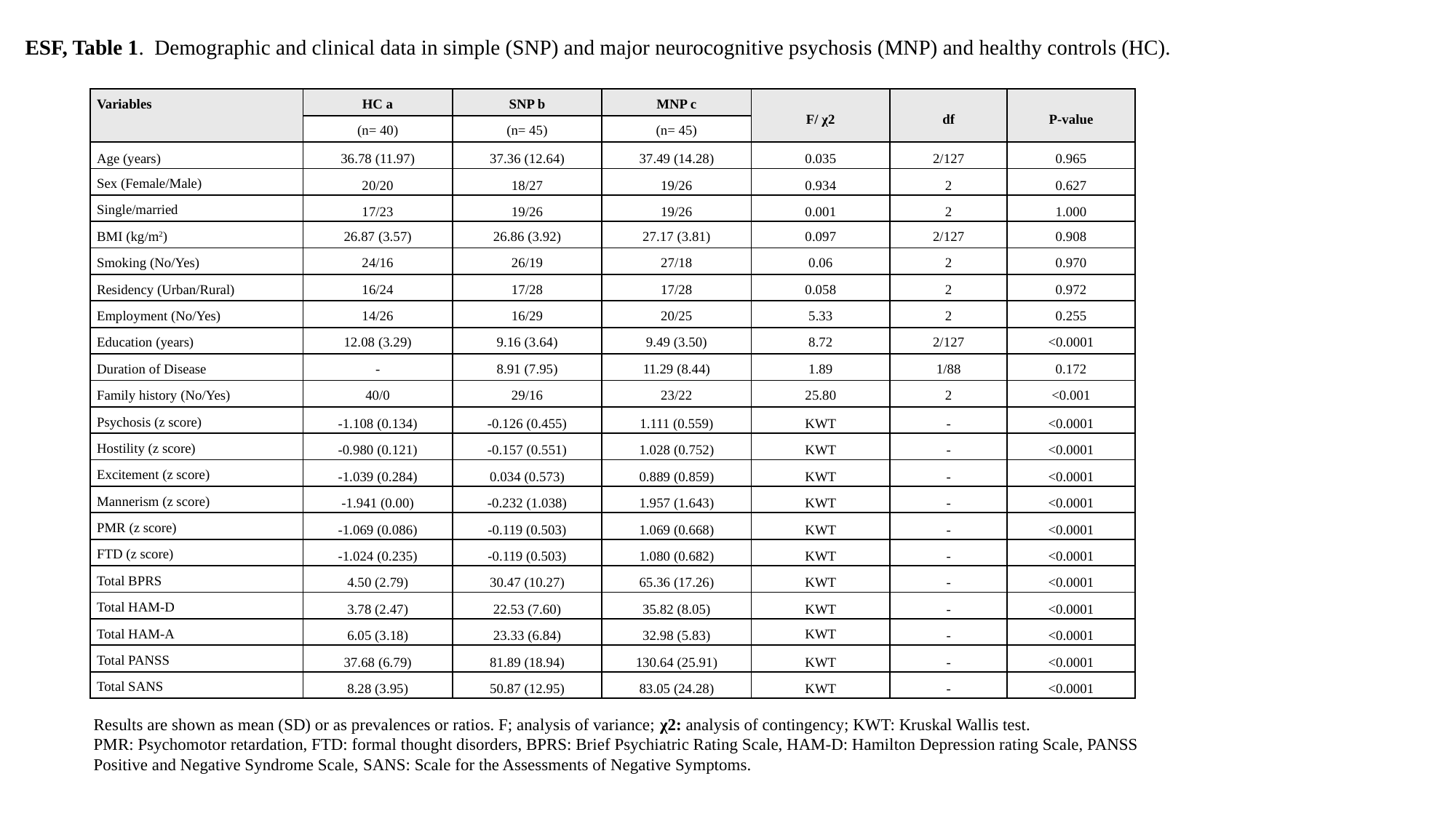

ESF, Table 1. Demographic and clinical data in simple (SNP) and major neurocognitive psychosis (MNP) and healthy controls (HC).
| Variables | HC a | SNP b | MNP c | F/ χ2 | df | P-value |
| --- | --- | --- | --- | --- | --- | --- |
| | (n= 40) | (n= 45) | (n= 45) | | | |
| Age (years) | 36.78 (11.97) | 37.36 (12.64) | 37.49 (14.28) | 0.035 | 2/127 | 0.965 |
| Sex (Female/Male) | 20/20 | 18/27 | 19/26 | 0.934 | 2 | 0.627 |
| Single/married | 17/23 | 19/26 | 19/26 | 0.001 | 2 | 1.000 |
| BMI (kg/m^2^) | 26.87 (3.57) | 26.86 (3.92) | 27.17 (3.81) | 0.097 | 2/127 | 0.908 |
| Smoking (No/Yes) | 24/16 | 26/19 | 27/18 | 0.06 | 2 | 0.970 |
| Residency (Urban/Rural) | 16/24 | 17/28 | 17/28 | 0.058 | 2 | 0.972 |
| Employment (No/Yes) | 14/26 | 16/29 | 20/25 | 5.33 | 2 | 0.255 |
| Education (years) | 12.08 (3.29) | 9.16 (3.64) | 9.49 (3.50) | 8.72 | 2/127 | <0.0001 |
| Duration of Disease | - | 8.91 (7.95) | 11.29 (8.44) | 1.89 | 1/88 | 0.172 |
| Family history (No/Yes) | 40/0 | 29/16 | 23/22 | 25.80 | 2 | <0.001 |
| Psychosis (z score) | -1.108 (0.134) | -0.126 (0.455) | 1.111 (0.559) | KWT | - | <0.0001 |
| Hostility (z score) | -0.980 (0.121) | -0.157 (0.551) | 1.028 (0.752) | KWT | - | <0.0001 |
| Excitement (z score) | -1.039 (0.284) | 0.034 (0.573) | 0.889 (0.859) | KWT | - | <0.0001 |
| Mannerism (z score) | -1.941 (0.00) | -0.232 (1.038) | 1.957 (1.643) | KWT | - | <0.0001 |
| PMR (z score) | -1.069 (0.086) | -0.119 (0.503) | 1.069 (0.668) | KWT | - | <0.0001 |
| FTD (z score) | -1.024 (0.235) | -0.119 (0.503) | 1.080 (0.682) | KWT | - | <0.0001 |
| Total BPRS | 4.50 (2.79) | 30.47 (10.27) | 65.36 (17.26) | KWT | - | <0.0001 |
| Total HAM-D | 3.78 (2.47) | 22.53 (7.60) | 35.82 (8.05) | KWT | - | <0.0001 |
| Total HAM-A | 6.05 (3.18) | 23.33 (6.84) | 32.98 (5.83) | KWT | - | <0.0001 |
| Total PANSS | 37.68 (6.79) | 81.89 (18.94) | 130.64 (25.91) | KWT | - | <0.0001 |
| Total SANS | 8.28 (3.95) | 50.87 (12.95) | 83.05 (24.28) | KWT | - | <0.0001 |
Results are shown as mean (SD) or as prevalences or ratios. F; analysis of variance; χ2: analysis of contingency; KWT: Kruskal Wallis test.
PMR: Psychomotor retardation, FTD: formal thought disorders, BPRS: Brief Psychiatric Rating Scale, HAM-D: Hamilton Depression rating Scale, PANSS
Positive and Negative Syndrome Scale, SANS: Scale for the Assessments of Negative Symptoms.

### Slide 3
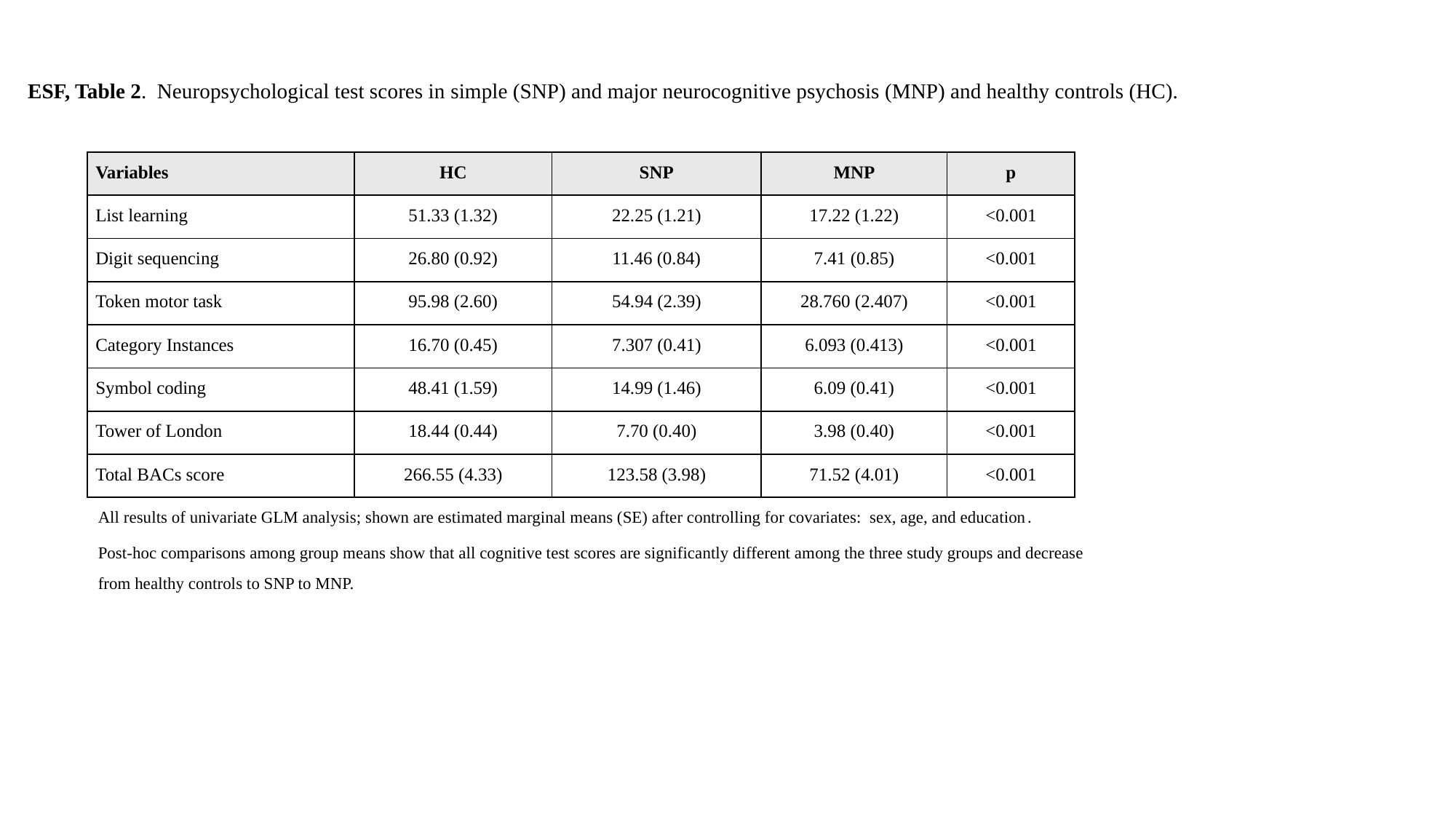

ESF, Table 2. Neuropsychological test scores in simple (SNP) and major neurocognitive psychosis (MNP) and healthy controls (HC).
| Variables | HC | SNP | MNP | p |
| --- | --- | --- | --- | --- |
| List learning | 51.33 (1.32) | 22.25 (1.21) | 17.22 (1.22) | <0.001 |
| Digit sequencing | 26.80 (0.92) | 11.46 (0.84) | 7.41 (0.85) | <0.001 |
| Token motor task | 95.98 (2.60) | 54.94 (2.39) | 28.760 (2.407) | <0.001 |
| Category Instances | 16.70 (0.45) | 7.307 (0.41) | 6.093 (0.413) | <0.001 |
| Symbol coding | 48.41 (1.59) | 14.99 (1.46) | 6.09 (0.41) | <0.001 |
| Tower of London | 18.44 (0.44) | 7.70 (0.40) | 3.98 (0.40) | <0.001 |
| Total BACs score | 266.55 (4.33) | 123.58 (3.98) | 71.52 (4.01) | <0.001 |
All results of univariate GLM analysis; shown are estimated marginal means (SE) after controlling for covariates: sex, age, and education.
Post-hoc comparisons among group means show that all cognitive test scores are significantly different among the three study groups and decrease from healthy controls to SNP to MNP.

### Slide 4
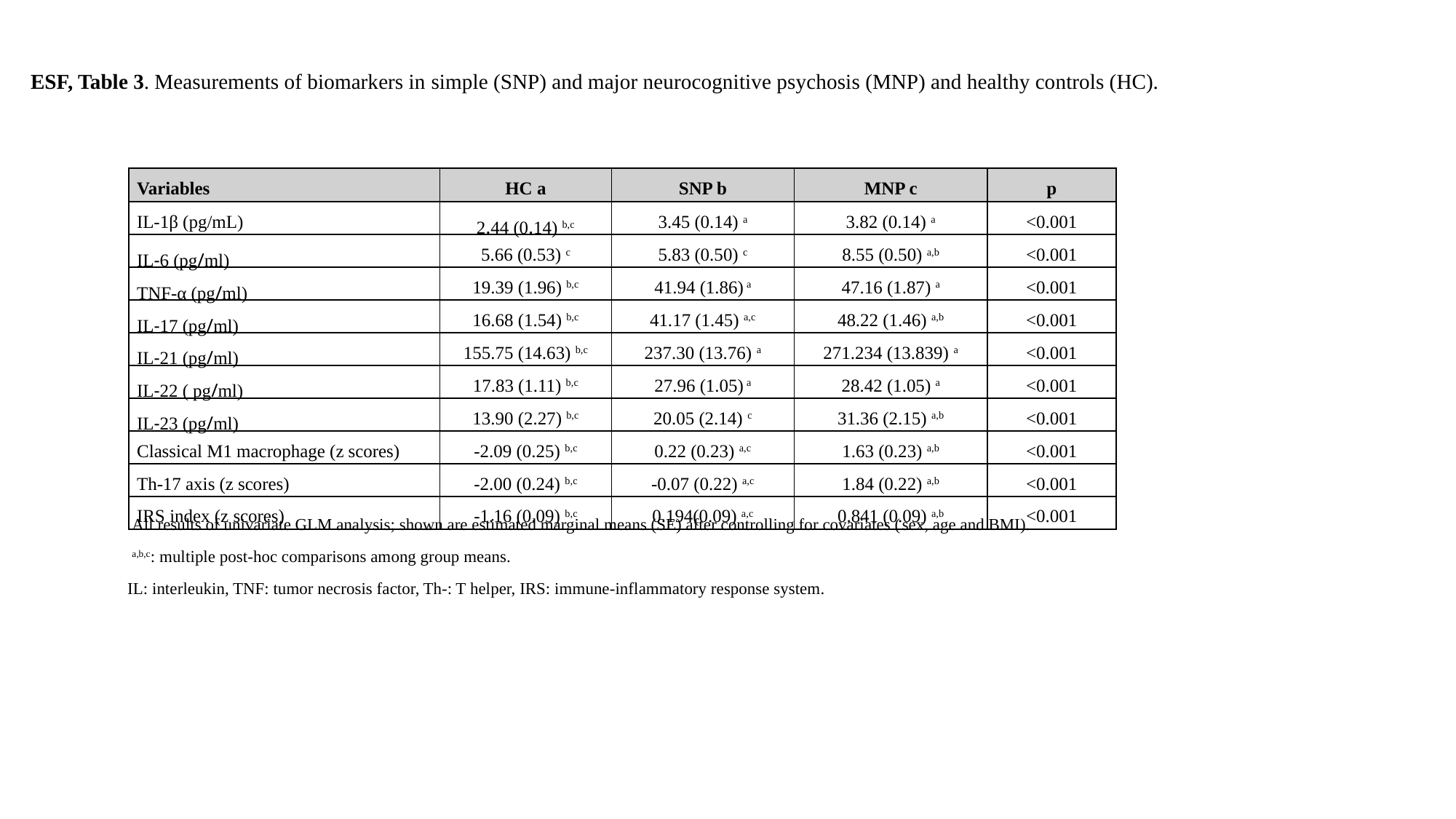

ESF, Table 3. Measurements of biomarkers in simple (SNP) and major neurocognitive psychosis (MNP) and healthy controls (HC).
| Variables | HC a | SNP b | MNP c | p |
| --- | --- | --- | --- | --- |
| IL-1β (pg/mL) | 2.44 (0.14) ^b,c^ | 3.45 (0.14) ^a^ | 3.82 (0.14) ^a^ | <0.001 |
| IL-6 (pg/ml) | 5.66 (0.53) ^c^ | 5.83 (0.50) ^c^ | 8.55 (0.50) ^a,b^ | <0.001 |
| TNF-α (pg/ml) | 19.39 (1.96) ^b,c^ | 41.94 (1.86) ^a^ | 47.16 (1.87) ^a^ | <0.001 |
| IL-17 (pg/ml) | 16.68 (1.54) ^b,c^ | 41.17 (1.45) ^a,c^ | 48.22 (1.46) ^a,b^ | <0.001 |
| IL-21 (pg/ml) | 155.75 (14.63) ^b,c^ | 237.30 (13.76) ^a^ | 271.234 (13.839) ^a^ | <0.001 |
| IL-22 ( pg/ml) | 17.83 (1.11) ^b,c^ | 27.96 (1.05) ^a^ | 28.42 (1.05) ^a^ | <0.001 |
| IL-23 (pg/ml) | 13.90 (2.27) ^b,c^ | 20.05 (2.14) ^c^ | 31.36 (2.15) ^a,b^ | <0.001 |
| Classical M1 macrophage (z scores) | -2.09 (0.25) ^b,c^ | 0.22 (0.23) ^a,c^ | 1.63 (0.23) ^a,b^ | <0.001 |
| Th-17 axis (z scores) | -2.00 (0.24) ^b,c^ | -0.07 (0.22) ^a,c^ | 1.84 (0.22) ^a,b^ | <0.001 |
| IRS index (z scores) | -1.16 (0.09) ^b,c^ | 0.194(0.09) ^a,c^ | 0.841 (0.09) ^a,b^ | <0.001 |
All results of univariate GLM analysis; shown are estimated marginal means (SE) after controlling for covariates (sex, age and BMI).
^a,b,c^: multiple post-hoc comparisons among group means.
IL: interleukin, TNF: tumor necrosis factor, Th-: T helper, IRS: immune-inflammatory response system.

### Slide 5
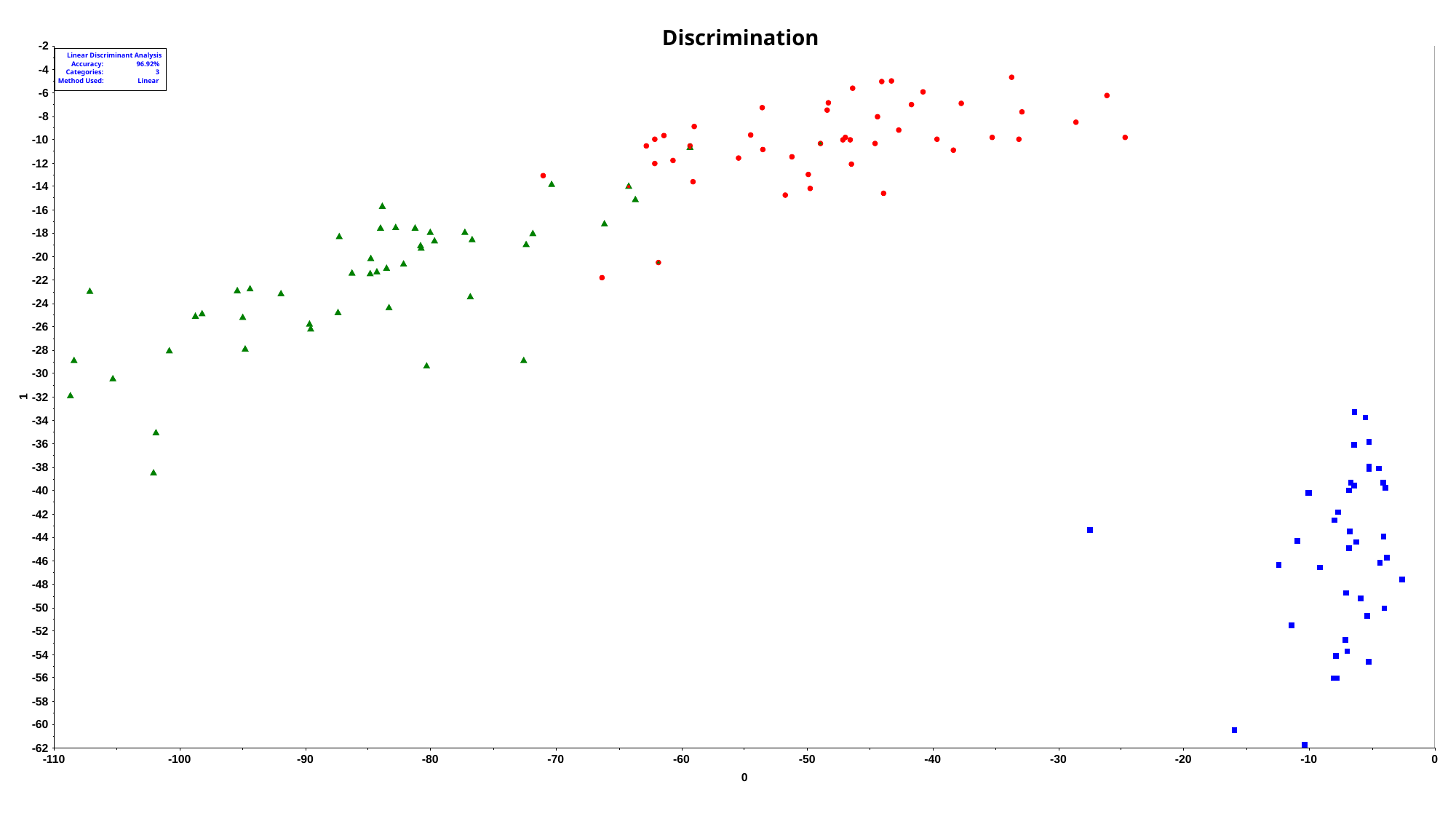

### Slide 6
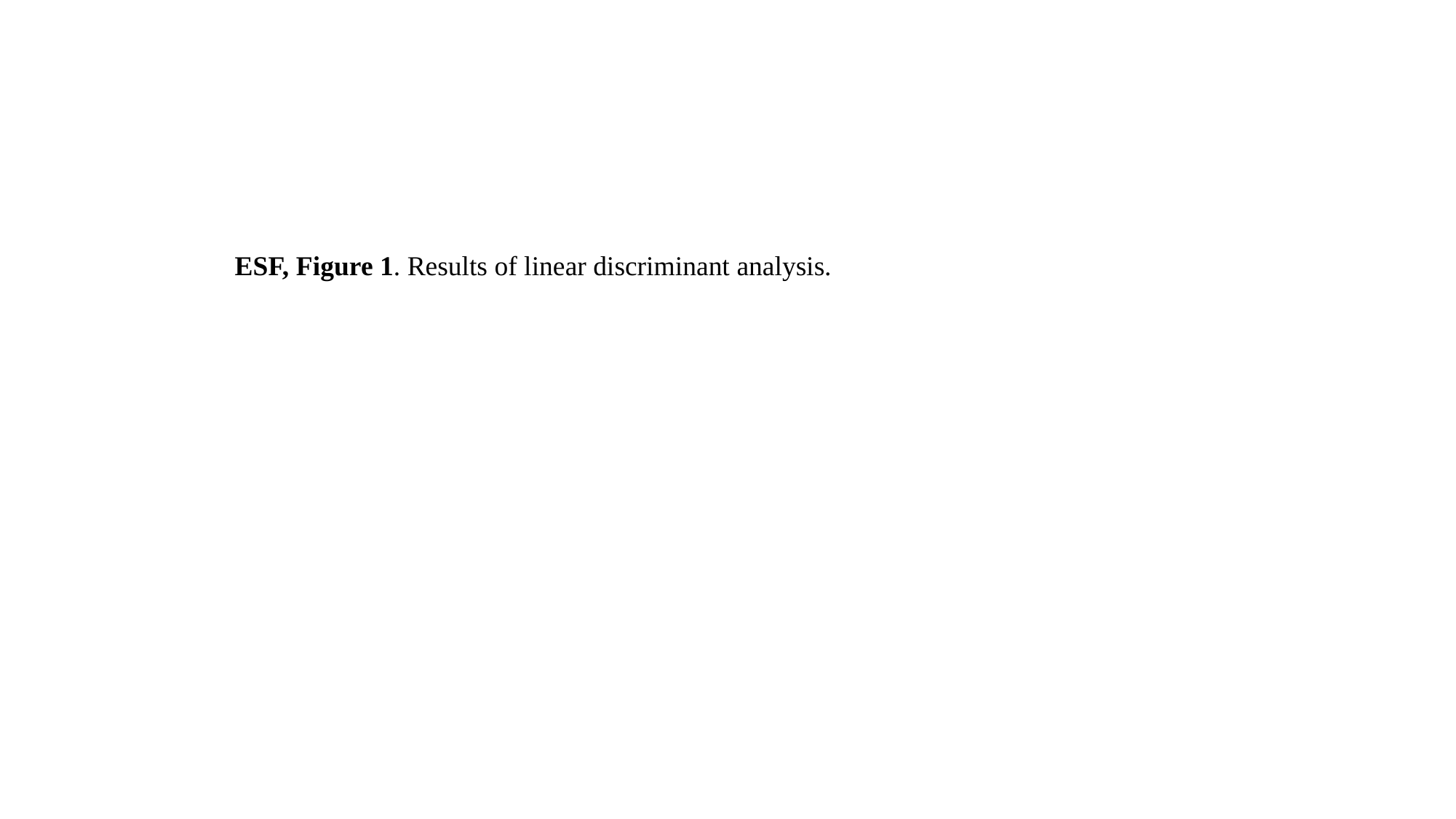

ESF, Figure 1. Results of linear discriminant analysis.

### Slide 7
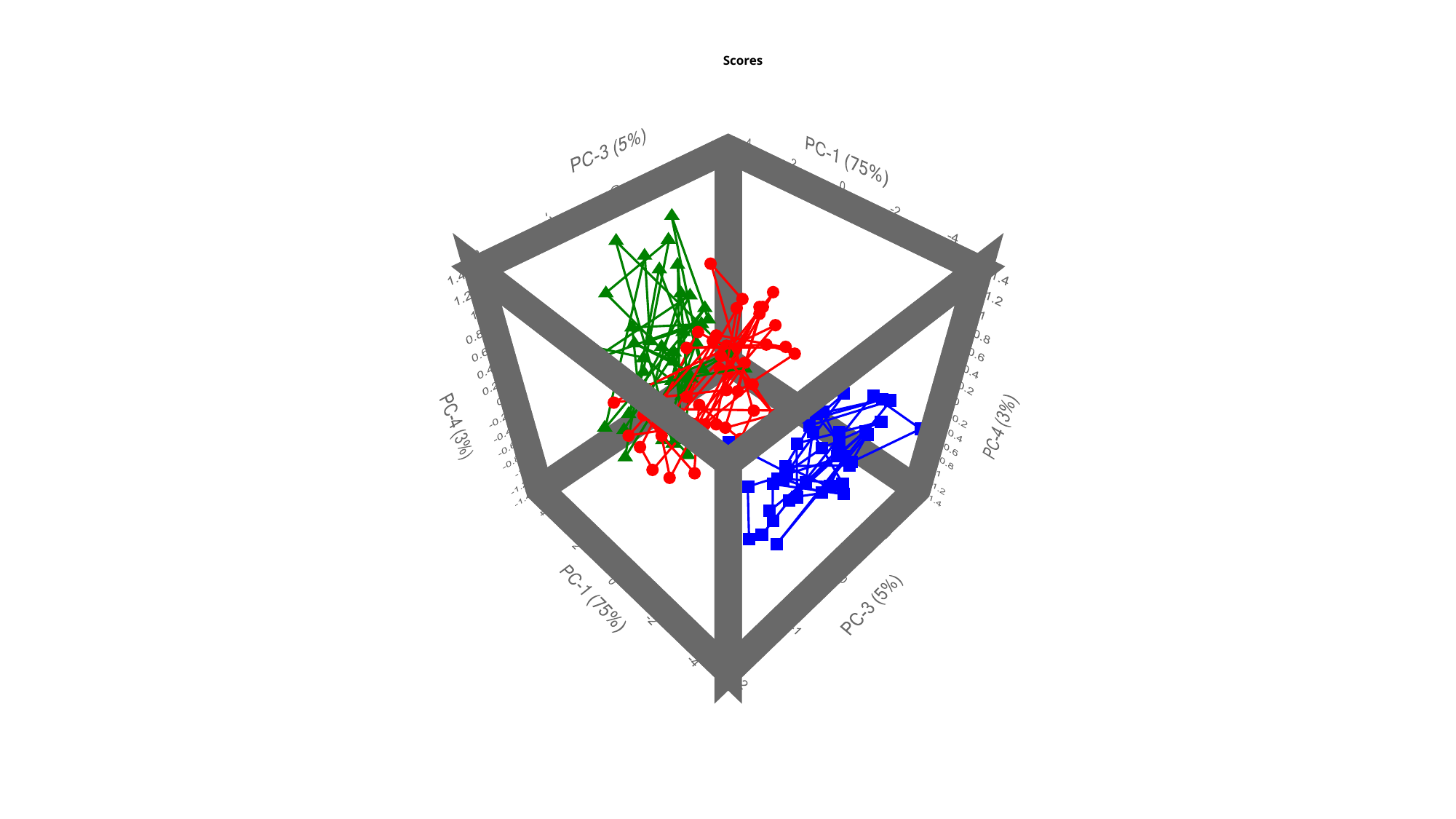

### Slide 8
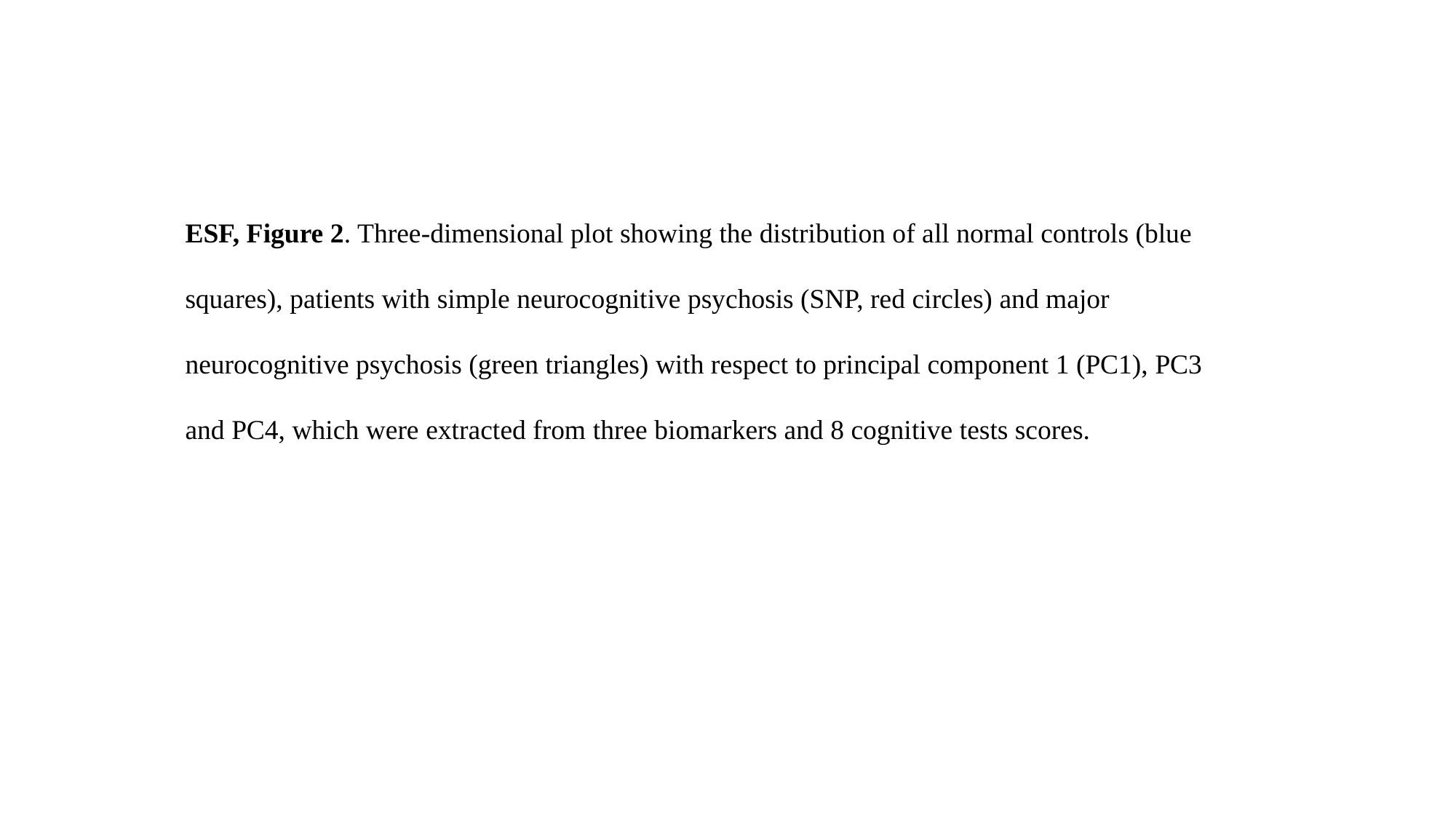

ESF, Figure 2. Three-dimensional plot showing the distribution of all normal controls (blue squares), patients with simple neurocognitive psychosis (SNP, red circles) and major neurocognitive psychosis (green triangles) with respect to principal component 1 (PC1), PC3 and PC4, which were extracted from three biomarkers and 8 cognitive tests scores.
